# Signatures of immune senescence predict outcomes and define checkpoint blockade-unresponsive microenvironments in acute myeloid leukemia

**DOI:** 10.1101/2022.02.08.22270578

**Authors:** Sergio Rutella, Jayakumar Vadakekolathu, Francesco Mazziotta, Stephen Reeder, Tung-On Yau, Rupkatha Mukhopadhyay, Benjamin Dickins, Heidi Altmann, Michael Kramer, Hanna Knaus, Bruce R. Blazar, Vedran Radojcic, Joshua F. Zeidner, Andrea Arruda, Mark D. Minden, Sarah K. Tasian, Martin Bornhäuser, Ivana Gojo, Leo Luznik

**Author notes:** Correspondence (S.R.), (L.L.). Equally contributing authors (Sergio Rutella, Ivana Gojo).

## Abstract

The function of senescent-like T cells, transcriptomic features of immune effector senescence (IES) and their influence on therapeutic response were investigated in independent AML clinical cohorts comprising 1,864 patients treated with chemotherapy and/or immune checkpoint blockade (ICB). We show that senescent-like bone marrow CD8^+^ T cells are impaired in killing autologous AML blasts, and that their proportion negatively correlates with overall survival (OS). We define new IES signatures using two gene expression platforms and report that IES scores correlate with adverse-risk molecular lesions, stemness, and poor outcomes as a potentially more powerful predictor of OS than 2017-ELN risk or LSC17 stemness score. IES expression signatures also identify an ICB- unresponsive tumor microenvironment and predict significantly worse OS in AML as well as in solid tumors. The newly described IES scores provide improved AML risk stratification and could facilitate the delivery of personalized immunotherapies to patients who are most likely to benefit.

## Introduction

Acute myeloid leukemia (AML) is a molecularly and clinically heterogeneous disease (Dohner et al., 2017). We recently identified bone marrow (BM) microenvironmental transcriptomic profiles that stratify patients with newly diagnosed AML into an immune-infiltrated and an immune-depleted subtype and that refine the accuracy of overall survival (OS) prediction beyond that afforded by current prognosticators (Vadakekolathu et al., 2020b). Several aspects of T-cell derangement affect AML response to standard-of-care chemotherapy, molecularly targeted therapies, and immunotherapies (Daver et al., 2019; Mathew et al., 2018; Vadakekolathu et al., 2020b; Vago and Gojo, 2020; Zeidner et al., 2020; Zeidner et al., 2021). In this respect, interferon (IFN)-γ-related RNA profiles in baseline BM samples predict response of chemotherapy-refractory AML to CD123×CD3 bispecific molecules (Uy et al., 2021; Vadakekolathu et al., 2020b).

The degree of cytotoxic CD8^+^ T-cell infiltration has been shown to correlate inversely with OS in select tumor types, including AML, because of the establishment of highly dysfunctional T-cell states (Geissler et al., 2015; Vadakekolathu et al., 2020b). Phenotypic and transcriptomic analyses have shown that CD8^+^ T cells from patients with AML exhibit features of exhaustion and senescence, and have identified a gene signature that diverges between chemotherapy responders and non-responders, with the former exhibiting upregulation of costimulatory and downregulation of apoptotic and coinhibitory T-cell signaling pathways (Knaus et al., 2018).

Exhaustion and senescence are dominant dysfunctional states of effector T cells that are increasingly recognized as major hurdles for success of cancer immunotherapy (Liu et al., 2020; Philip and Schietinger, 2021). Senescence and exhaustion share properties, but they may be functionally divergent (Akbar and Henson, 2011). Exhausted T cells express inhibitory receptors, including *PDCD1* (encoding PD-1), *CTLA4*, *HAVCR2* (encoding *TIM3*), *CD160* and *2B4* (encoding CD244), and display an impaired ability to secrete effector cytokines and to exert cytotoxic functions. Senescent T cells downregulate costimulatory molecules CD27 and CD28, express senescence-associated surface markers B3GAT1 and KLRG1, as well as MAPK p38 and γH2AX intracellular molecules, remain metabolically active and continue to secrete pro-inflammatory cytokines (Henson et al., 2014; Liu et al., 2021), but their cytotoxic anti-tumor activity is unclear. Whilst more is known about the role of T-cell exhaustion in immunotherapy responses, the contribution of T-cell senescence to anti-cancer immunity is unknown (Akbar and Henson, 2011).

In the current study, we characterize how leukemia promotes the generation of senescent-like CD8^+^ T cells and their prognostic relevance in patients with AML. We hypothesize that elucidation of an immune senescence transcriptional signature in the BM of newly diagnosed AML patients could identify individuals more likely to respond to immunotherapy and predict outcomes. We generated RNA expression datasets from patients with AML treated with conventional cytotoxic chemotherapy or with the hypomethylating agent, azacitidine, in combination with immune checkpoint blockade (ICB) with pembrolizumab (monoclonal antibody targeting PD-1; AZA+Pembro). We integrated these with publicly available gene expression data from multiple cohorts of children and adults with AML to validate our RNA metric of immune effector senescence (IES) and we analyzed BM samples collected longitudinally at time of AML onset and response assessment (**Figure S1**). The derived gene signatures of IES correlated with molecular features of stemness and with distinct clinical characteristics. IES signatures served as a reliable biomarker to stratify OS after standard-of-care therapy and immune checkpoint blockade (ICB), both in AML and in melanoma (a paradigm for successful immunotherapy actualization) (Larkin et al., 2019; Robert et al., 2020).

## Results

### Functional and transcriptional signature of immune senescence in AML

AML blasts are known to be an extrinsic modifier of T-cell responses (Deng et al., 2018; Mussai et al., 2013; Pyzer et al., 2017; Williams et al., 2019). Initially, we aimed to evaluate experimentally whether AML blasts impact T-cell proliferation, activation, and expression of phenotypic markers of senescence through direct contact or by secreting soluble mediators. Flow cytometry-sorted BM T cells and AML blasts from newly diagnosed patients were co-cultured, either in direct contact or separated by transwell inserts, and stimulated as previously described (Knaus et al., 2018). We found that AML blasts induced expression of two well-characterized senescence markers, CD57 and γH2AX, on AML CD8^+^ T cells in both experimental conditions. Consistent with previous observations (Knaus et al., 2018), direct contact of AML blasts with T cells resulted in decreased expression of activation/proliferation CD25, ICOS, and Ki-67 (**Figure 1A**). However, when T cells were separated from AML blasts by a transwell, the expression of activation markers and Ki-67 equaled that on CD8^+^ T cells stimulated in the absence of AML blasts. These findings suggest that interactions between leukemia blasts and T cells occurring in the local milieu impair T-cell activation through direct contact, while promotion of senescence occurs primarily through bystander modulation. These effects seem to be AML blast-specific since co-culture with healthy donor monocytes did not affect any of the markers examined (**Figure 1B**).

**Figure 1.**
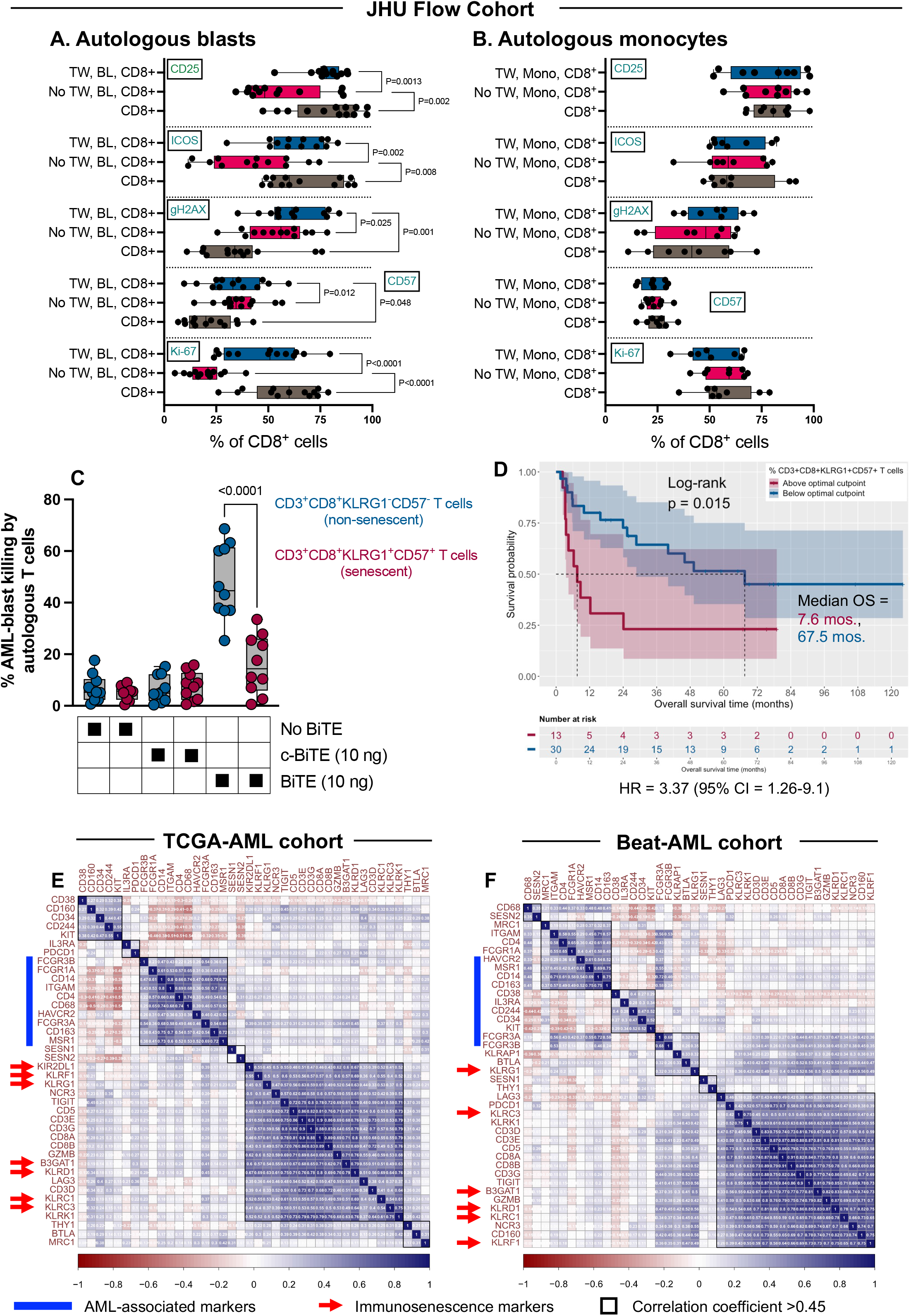
Markers of immune effector senescence (IES) correlate with impaired T-cell killing and with poor clinical outcomes. (A) Flow-sorted AML blasts were co-cultured with autologous, patient-derived CD8^+^ T cells for 5 days. Data were compared using the Wilcoxon matched pairs signed rank test. TW = transwell insert; BL = AML blasts; Mono = monocytes. (B) Flow-sorted monocytes were co-cultured with autologous, patient-derived CD8^+^ T cells for 5 days. Data were compared using the Wilcoxon matched pairs signed rank test. TW = transwell insert; BL = AML blasts; Mono = monocytes. (C) *In vitro* killing of primary AML blasts after 48-hour culture with autologous T cells in the presence of anti–CD33/CD3 and control bi-specific T-cell engager (BiTE) antibody constructs (10 ng/mL). Bone marrow T cells were flow-sorted into a senescent CD3^+^CD8^+^CD57^+^KLRG1^+^ and a non-senescent CD3^+^CD8^+^CD57^-^KLRG1^-^ subpopulation before being co-cultured with CD33^+^CD34^+^ AML blasts from the same patient (effector/target ratio = 1:5; BM samples from 10 patients in the JHU1 cohort were used for these experiments). T-cell cytotoxicity against CD33^+^CD34^+^ primary AML cells was determined by flow cytometry, as detailed in the STAR Methods section. (D) Kaplan-Meier estimates of overall survival (OS) in patients (JHU1 cohort) with senescent CD3^+^CD8^+^CD57^+^KLRG1^+^ T cells above (magenta line) and below (blue line) the optimal cut- point, which was computed using the *maxstat* package in R. Survival curves were compared using a log-rank test (*survminer* package in R). Median OS is indicated (color-coded by the optimal cut-point of the proportion of CD3^+^CD8^+^CD57^+^KLRG1^+^ T cells). HR = hazard ratio; CI = confidence interval. (E-F) Correlograms showing co-expression of NK-related antigens and T-cell markers (red arrows), including *CD8A*, *CD8B*, *CD3E*, *CD3G* and *CD5* (*corrplot* package in R) in TCGA- AML and Beat-AML cases. Data were retrieved through the cBioPortal for Cancer Genomics (https://www.cbioportal.org/) and the Vizome portal (http://www.vizome.org/aml/), respectively (Tyner et al., 2018). NK-cell, T-cell, monocyte-macrophage (*CD14*, *CD68*, *CD163*) and AML-associated markers (*CD34*, *IL3RA*, *KIT*, *THY1*) were selected by integrating knowledge from multiple publications (Bagaev et al., 2021; Ehninger et al., 2014; Knaus et al., 2018).

Given the high frequency of senescent-like T cells in the BM of patients with AML (Knaus et al., 2018), we next investigated *in vitro* cytotoxicity of flow-sorted, BM-derived senescent (CD3^+^CD8^+^CD57^+^KLRG1^+^) and non-senescent (CD3^+^CD8^+^CD57^-^KLRG1^-^) T cells against autologous AML blasts using an anti–CD3/CD33 bispecific T-cell engaging (BiTE) antibody construct (Krupka et al., 2014; Laszlo et al., 2014). As shown in **Figure 1C**, senescent-like T cells were significantly impaired in their ability to lyse AML blasts compared with their non-senescent counterpart. These findings could explain the inferior killing ability of CD3/CD33 BiTE construct when using patient T cells versus those of healthy controls (Harrington et al., 2015). Analysis of 43 patients with newly diagnosed AML (JHU1 cohort; **Table S1**) also revealed that a higher proportion of senescent-like (CD3^+^CD8^+^CD57^+^KLRG1^+^) T cells in baseline BM samples was associated with significantly worse OS (*P*=0.004) after treatment with standard chemotherapy (**Figure 1D**).

Transcriptional profiling of the tumor microenvironment (TME) has been used to identify immunological signatures and biological processes and to develop predictors of protective immunity (Bagaev et al., 2021; Thorsson et al., 2018). We therefore sought to derive gene expression signatures of T-cell immunosenescence in the AML BM microenvironment. We compiled a manually curated IS gene signature, which encompassed *KLRG1*, *CD57* and other senescence markers (*KLRC1*, *KLRC3*, *KLRD1*, *KLRF1*, and *CD158A*) previously shown to be expressed by circulating CD8^+^ T cells from patients with AML (Knaus et al., 2018). We used RNA-seq data and related clinical information from the TCGA-AML and Beat-AML Master Trial (hereafter Beat-AML) cohorts (n=147 and n=264 unique patients, respectively) and correlated the expression of genes in the IS signature with markers of immune cells and leukemia blasts.

Pearson correlation analysis showed a positive correlation between IS genes and T-cell markers, but not with markers of AML blasts (*CD34*, *CD38*, *IL3RA*, *KIT*) nor with markers of accessory cells of the monocyte/macrophage lineage (*CD14*, *CD68*, *CD163*; **Figure 1E-1F**). Interestingly, the IS genes clustered together with inhibitory receptors (*TIGIT*, *LAG3*, *CD160*, *PDCD1*), NK and T-cell markers, an observation that is in line with recent studies showing the upregulation of NK antigens on senescent-like T cells (Pereira et al., 2020) and on dysfunctional chimeric antigen receptor (CAR) T cells (Good et al., 2021). The clustering of T-cell exhaustion and senescence genes is consistent with our previous flow cytometry studies (Knaus et al., 2018), suggesting that T cells in the AML microenvironment exhibit features of both biological processes (Akbar and Henson, 2011). Overall, the above findings indicate that both cellular and transcriptional signatures of CD8^+^ T-cell senescence are present in newly diagnosed AML patients, and that the abundance of senescent-like T cells may correlate with anti-leukemia responses and OS after induction chemotherapy.

### Identification of a BM immune effector senescence (IES) signature in two discovery AML cohorts

We hypothesized that probing Immune Signature Data Base (ImmuneSigDB) gene sets might reveal core biological processes involved in anti-tumor immune responses and in therapeutic outcomes. To this end, both TCGA-AML and Beat-AML cases (**Table S1**) were split into quartiles based on average expression levels of the seven immunosenescence genes. Gene set enrichment analysis (GSEA) was used to identify core gene sets accounting for the enrichment signal in immunosenescence^high^ (highest quartile) versus immunosenescence^low^ cases (lowest quartile; **Figure 2A**). Among the 4,872 curated gene sets from the ImmuneSigDB, only gene sets with false discovery rate (FDR)<0.05 and normalized enrichment score (NES)>2.0 (n=123 and n=126 gene sets at the intersection of TCGA-AML and Beat-AML cases, respectively) were carried forward for leading-edge analysis. This analysis identified 172 biologically related genes that are common to multiple significantly enriched ImmuneSigDB gene sets and that contribute most to the enrichment signal (**Figure S2A** and **Table S2**). The uniform manifold approximation and projection (UMAP) of single-cell RNAseq data from patients with AML (Dufva et al., 2020) revealed that single-cell clusters assigned to CD8^+^ T cells, CD4^+^ T cells and NK cells were highly enriched in this signature (**Figure S2B**). Features of cellular senescence are manifested by T cells in all differentiation states (Martinez-Zamudio et al., 2021; Pereira et al., 2020), and the 172 genes showed broad transcriptional overlap among multiple effector subsets. We hereafter refer to this gene set as the immune effector senescence (IES172) signature.

**Figure 2.**
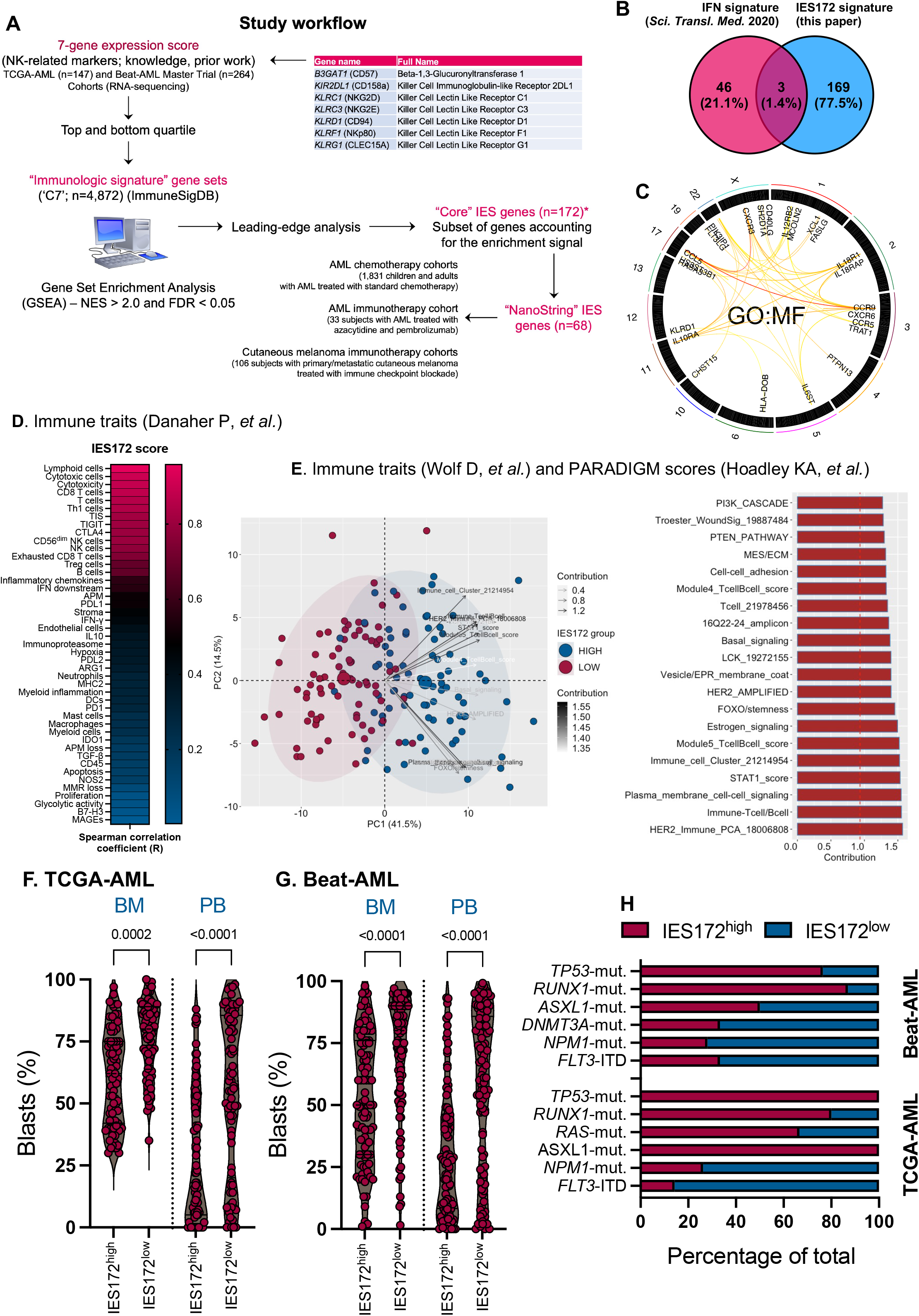
Signatures of immune effector senescence (IES) correlate with immune infiltration and with adverse-risk molecular features in the TCGA-AML and Beat-AML cohorts. (A) Study workflow. ImmuneSigDB = Immune Signature Data Base, a collection of approximately 5,000 annotated sets of up- or down-regulated genes pertaining to immune biology (Godec et al., 2016); IES = immune effector senescence; NES = normalized enrichment score; FDR = false discovery rate. (B) Overlap between the IES172 signature genes from this study and published signatures that predict chemotherapy refractoriness as well as response to bispecific T-cell engagers (Vadakekolathu et al., 2020b). The Venn diagram was generated using the *ggVennDiagram* package in R (Gao et al., 2021). IFN = interferon. (C) Semantic similarity between the IES172 genes in the context of their chromosomal location. The circular plot was drawn using the *XGR* (eXploring Genomic Relations) web tool (Fang et al., 2016) and the IES172 genes as input. The degree of similarity between genes is visualized by the color of links. GO:MF = gene ontology molecular functions. (D) Correlation between the IES172 score and previously published immune traits (n=45) (Danaher et al., 2017; Vadakekolathu et al., 2020b). Signature scores are available through the original publications. (E) Correlation between the IES172 score and previously published immune traits and PARADIGM scores (n=68; Wolf D, *et al*.), which were downloaded from the UCSC Xena data portal (https://xenabrowser.net/datapages/) (Goldman et al., 2020; Vaske et al., 2010). The principal component analysis (PCA) plot was generated using the *ggplot2* R package. The top contributors to the first and second PC (n=20) are shown as a bar graph. The dotted reference line indicates the expected value if the contribution were uniform. Any feature above the reference line can be considered as important in contributing to the dimension. (F) IES172 scores and leukemia burden (percentage of blasts) at diagnosis in TCGA-AML cases. Data were compared using the Mann Whitney *U* test for unpaired determinations. BM = bone marrow; PB = peripheral blood. (G) IES172 scores and leukemia burden (percentage of blasts) at diagnosis in Beat-AML cases. Data were compared using the Mann Whitney *U* test for unpaired determinations. (H) Stacked bar graph showing the proportion of IES172^high^ and IES172^low^ cases harboring mutations of *TP53*, *RUNX1*, *ASXL1*, *DNMT3A*, *NPM1* and *FLT3*-internal tandem duplication (ITD). Mut. = mutated.

The IES172 genes showed minimal overlap with knowledge-based transcriptional signatures of T-cell exhaustion, CAR T-cell dysfunction and solid tumor response to ICB (**Figure S2C**) (Good et al., 2021; Gueguen et al., 2021; Guo et al., 2018) and with IFN-related RNA profiles carrying prognostic significance in AML (**Figure 2B**) (Vadakekolathu et al., 2020b). The semantic similarity between IES172 genes in the context of their chromosomal location is shown in **Figure 2C**. No genes in the IES172 signature were on chromosome 7, the loss of which has been associated with failure to respond to PD1 blockade (Abbas et al., 2021). Furthermore, IES172 genes were enriched in Kyoto Encyclopedia of Genes and Genomes (KEGG) pathways related to T helper differentiation, T-cell receptor (TCR) signaling, and T/NK cell-mediated cytotoxicity, as well as miRNAs implicated in cancer immune escape and immune metabolism (Pu et al., 2021; Su et al., 2021; Vignard et al., 2020) (**Figure S2D-S2E** and **Table S3**). Using a broad collection of immune gene sets (Ayers et al., 2017; Campbell et al., 2018; Danaher et al., 2017; Goldman et al., 2020), we found that IES states correlate with lymphoid cells, CD8^+^ T-cell and NK-cell infiltration, the tumor inflammation signature (TIS) score and immune checkpoints *TIGIT*, *CTLA4* and *PD-L1* (**Figure 2D**). A principal component analysis (PCA) with publicly available immune signatures and PARADIGM integrated pathways (Hoadley et al., 2014; Vaske et al., 2010) as dependent variables lent further support to the association between IES states and immune infiltration and identified T-cell/B-cell scores, STAT1 signaling and stemness-related pathways as the top discriminative features (**Figure 2E**).

We looked for correlations between the IES172 score in diagnostic samples and pre-treatment variables in patients from the TCGA-AML and Beat-AML cohorts (**Table S4**) (Tyner et al., 2018). We found that the IES172 score does not correlate with patient age at diagnosis, 2017-ELN risk category or mutation count (**Figure S3A-S3B**) and was higher in AML cases with low leukemia burden (**Figure 2F-2G**) or harboring *TP53*, *RUNX1*, and *RAS* mutations (100%, 80% and 67% of cases, respectively; **Figure 2H**). These findings are consistent with previous reports on the immune landscape of *TP53*- and *RUNX1*-mutated AML (Sallman et al., 2020; Vadakekolathu et al., 2020a) and with the inverse correlation between immune infiltration and tumor purity (Miranda et al., 2019). The analysis of Beat- AML cases (n=264, of which 195 have chemotherapy response data) revealed significantly higher IES172 scores in patients with primary induction failure (PIF; n=63) compared with those achieving complete remission (CR; n=132; *P*=0.0044; **Figure S3C-S3D**). When analyzing matched samples collected at baseline and after induction chemotherapy (available only for 13 patients in the Beat-AML series), we found that the IES172 score was significantly higher in post-chemotherapy BM samples (*P*=0.0046; **Figure S3E**).

### IES scores correlate with transcriptomic features of AML stemness and stratify survival

The 17-gene leukemia stem cell (LSC17) score has previously been associated with poor clinical outcomes and with *TP53* and *RUNX1* mutational status in *de novo* AML (Bill et al., 2020; Ng et al., 2016). The LSC17 score discriminated survival outcomes in TCGA-AML and in Beat-AML patient cohorts (**Figure S4A-S4B**). The LSC17 score was not collinear with previously published immune cell type-specific gene signatures (Danaher et al., 2017), immune checkpoints, IFN-γ-related gene programs and the IES172 signature score (**Figure 3A**) (Ott et al., 2019) and was significantly higher in samples with above-median IES172 scores (**Figure 3B**). This finding was corroborated using xCELL, an ssGSEA-based tool that infers cellular content in the tumor microenvironment (**Figure 3C**) (Aran et al., 2017). When patients were stratified into IES172^high^ and IES172^low^, the LSC17 score retained its ability to predict overall survival (OS; **Figure 3D-3E**).

**Figure 3.**
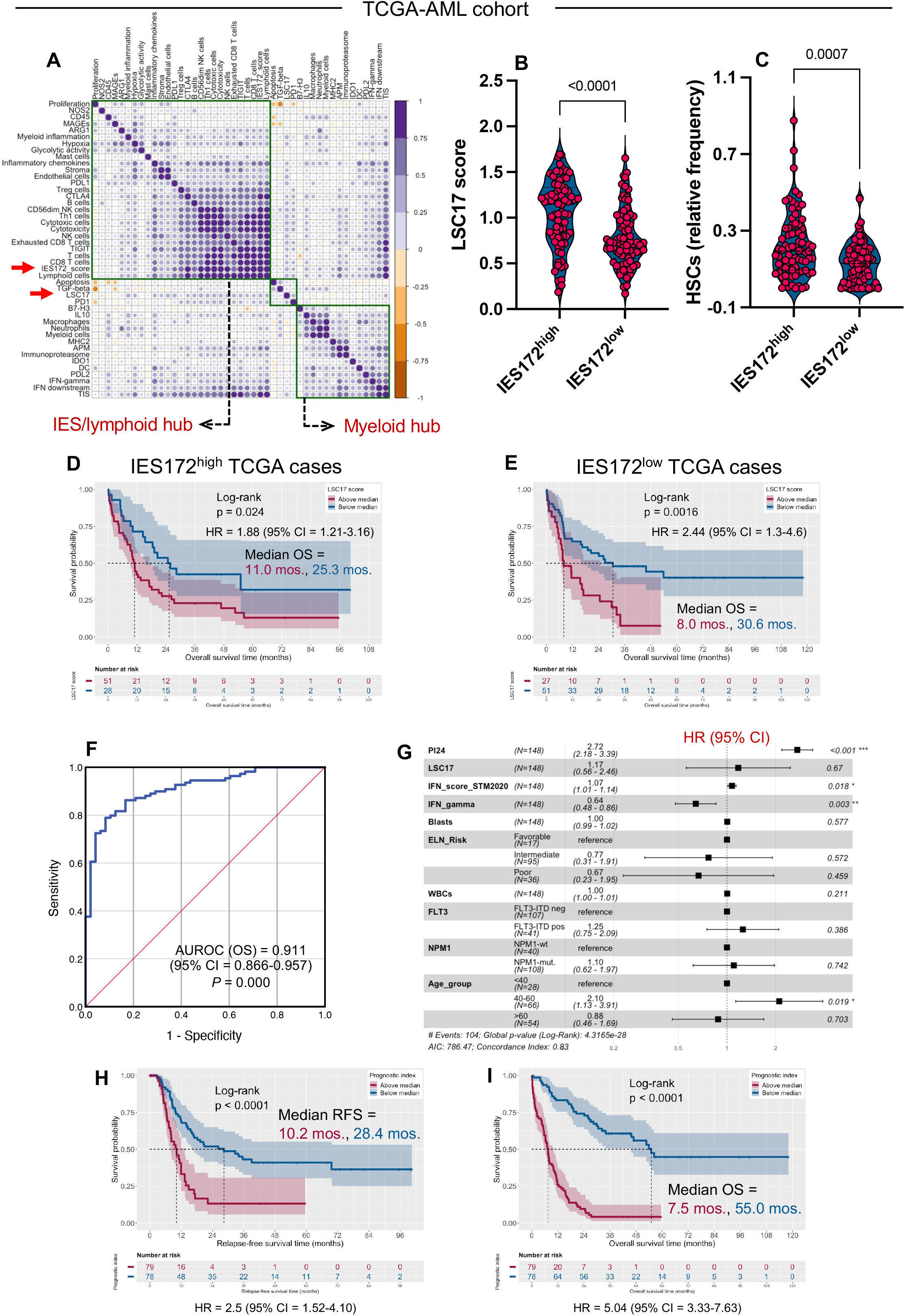
Co-varying gene programs of immune effector senescence (IES) and stemness in the TCGA-AML cohort. (A) Pairwise correlation between transcriptomic traits of immune infiltration and LSC17/IES172 scores (unsupervised hierarchical clustering). Non-significant *P* values on the correlation matrix plot are shown as blank boxes (*corrplot* package in R). Modules, including traits that are densely connected (hubs), are identified based on hierarchical clustering (Euclidean distance, complete linkage) and are shown in black boxes. The red arrows highlight the IES172 and LSC17 scores. (B) LSC17 score in patients categorized as IES172^high^ and IES172^low^ (median split). Data were compared using the Mann Whitney *U* test for unpaired determinations. (C) Relative frequency of hematopoietic stem cells (HSCs) in patients categorized as IES172^high^ and IES172^low^ (median split) as estimated by xCELL (Aran et al., 2017). Pre- calculated TCGA scores were downloaded from https://xcell.ucsf.edu/. (D) Kaplan-Meier estimates of overall survival (OS) in IES172^high^ patients with above median (magenta line) and below median (blue line) LSC17 scores. Survival curves were compared using a log-rank test (*survminer* package in R). HR = hazard ratio. (E) Kaplan-Meier estimates of overall survival (OS) in IES172^low^ patients with above median (magenta line) and below median (blue line) LSC17 scores. Survival curves were compared using a log-rank test (*survminer* package in R). (F) AUROC curve measuring the predictive ability of PI24 genes (blue curve) for OS. CI = confidence interval. AUROC = 1.0 denotes perfect prediction and AUROC = 0.5 denotes no predictive ability. (G) Forest plot (*ggforest* function in *survminer* package in R) of pre-treatment features (white blood cell count [WBC] at diagnosis, percentage of bone marrow blasts, *FLT3*-ITD and *NPM1* mutational status, patient age at diagnosis), and RNA-based scores associated with survival in multivariate Cox proportional hazard analyses [PI24, LSC17 and interferon (IFN) scores] (Ng et al., 2016; Vadakekolathu et al., 2020a; Vadakekolathu et al., 2020b). HR = hazard ratio for death. (H) Kaplan-Meier estimates of relapse-free survival (RFS) in TCGA-AML patients with above median (magenta line) and below median (blue line) PI24. The PI was calculated using β values from Cox regression analyses of gene expression and patient survival, as previously published (Wagner et al., 2019). Survival curves were compared using the log-rank test. (I) Kaplan-Meier estimates of OS in TCGA-AML patients with above median (magenta line) and below median (blue line) PI24. Survival curves were compared using the log-rank test.

To determine the parameters most predictive of outcomes in the IES172 signature, we used the statistical method LASSO to fit an L1-regularized liner model (Tibshirani, 1997) which revealed a parsimonious set of 24 genes predictive of OS. We then generated a prognostic index (PI) using β values from Cox regression analyses of gene expression and OS (**Figure S5**) (Wagner et al., 2019). The 24-gene PI was an independent predictor of OS with an AUROC value of 0.911 in the TCGA-AML cohort (**Figure 3F**). In multivariable analyses controlling for tumor purity, the 24-gene PI was a more powerful predictor of OS than the LSC17 score (Ng et al., 2016), the IFN-related score (Vadakekolathu et al., 2020b) and other established AML prognosticators, including *FLT3*-ITD and *NPM1* mutational status at diagnosis (**Figure 3G**). On stratifying patients above or below the median PI24, we found that subjects with above-median PI24 experienced significantly inferior RFS and OS (*P*<0.0001 for both; **Figure 3H-3I**). High PI24 scores were also associated with significantly inferior OS compared with patients with low PI24 in the Beat-AML cohort (*P*=0.012; **Figure S6A**). In agreement with TCGA data, the 24-gene PI was a good predictor of OS, with an AUROC value of 0.805 (**Figure S6B**).

As shown in **Figure S7A-S7D**, an optimal 24-gene PI cut-point of 1.73 parsed the TCGA population into subgroups with maximally different survival probabilities. Furthermore, patients in the highest quartile of PI24 values had dismal clinical outcomes (1-year RFS and OS rates of 0% and 3%, respectively) compared with patients in the lowest quartile (1-year RFS and OS rates of 74% and 97%, respectively). These findings were validated in the Beat-AML cohort (**Figure S7E-S7F**) and in another large cohort of 562 adult subjects with AML treated on the German AMLCG 1999 trial (GSE37642; **Figure S8A-8B**) (Herold et al., 2018).

### Validation of IES scores in relation to immune infiltration, stemness, chemotherapy refractoriness and patient outcome in independent AML cohorts

Benefiting from our previous work that harnessed large numbers of clinically annotated AML samples (Knaus et al., 2018; Vadakekolathu et al., 2020b) and with the aim to develop a gene expression assay that can be rapidly implemented in clinical practice, we turned to the nCounter platform (NanoString Technologies; Seattle, WA) (Jiang et al., 2018; Ng et al., 2016). We initially mined our published AML dataset (GSE134589; PMCC cohort encompassing 290 patients with newly diagnosed AML) and identified 68 genes that are shared between the RNA sequencing based IES172 signature and the NanoString panel genes (IES68; **Figure S5** and **Table S2**). Both the IES172 and the IES68 signatures showed enrichment in genes with annotated functions in cytokine/chemokine signaling, TCR signaling, co-stimulation by the CD28 family and PD-1/PD-L1 immune checkpoints in cancer (**Figure 4A**). Overlaying IES transcriptional signatures onto the UMAP of scRNA-seq data revealed that IES genes largely map to CD8^+^ T-cell and NK-cell clusters (**Figure 4B**). As shown in **Figure 4C-4D**, and in agreement with prior analyses, the IES68 signature was higher in tumors infiltrated with CD8^+^ and NK cells and characterized by the expression of inhibitory molecules, and inversely correlated with leukemia burden.

**Figure 4.**
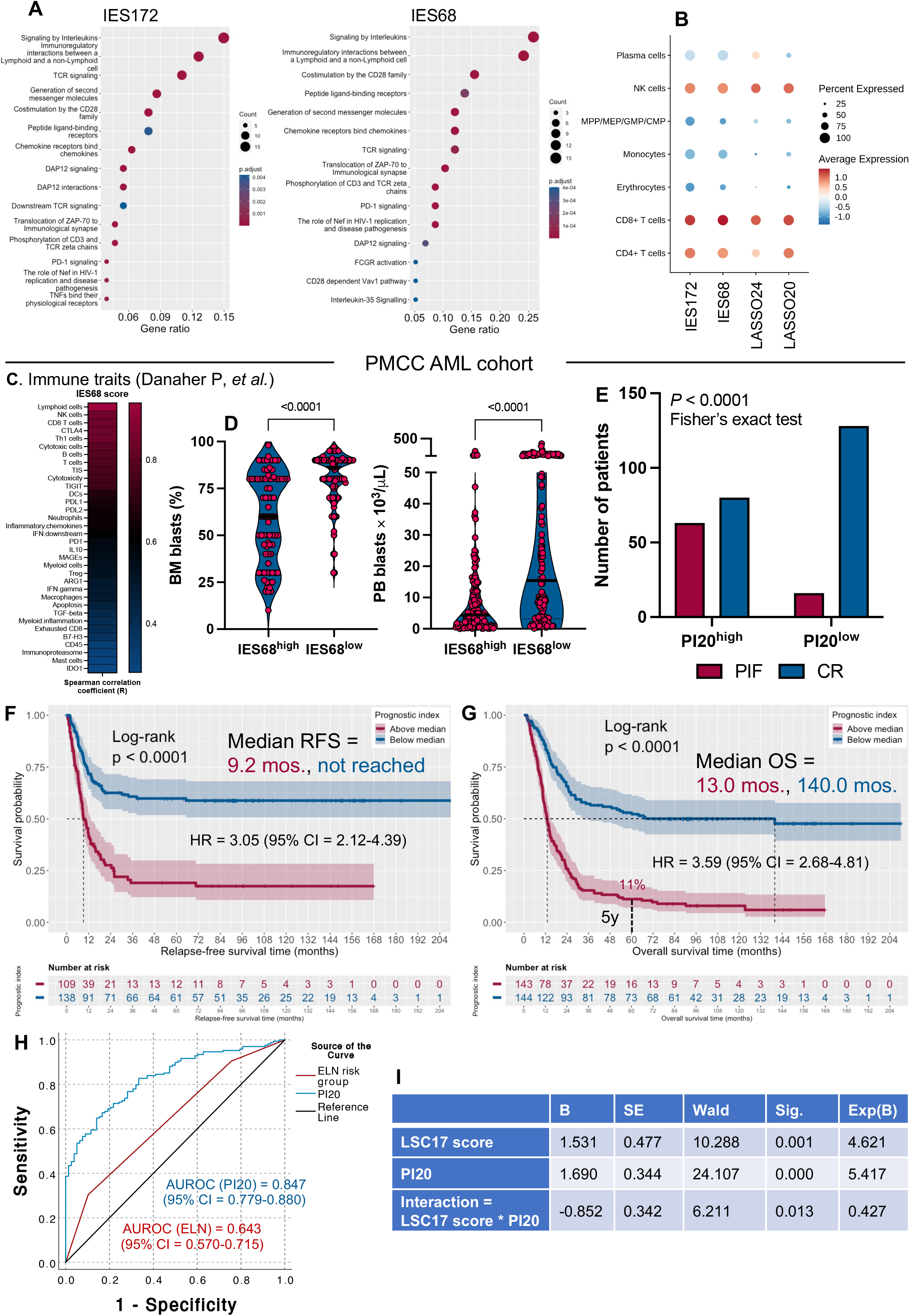
Immune effector senescence (IES) scores correlate with immune infiltration, stemness, primary induction failure and patient outcome in an external AML cohort. (A) Bubble plot depicting enriched REACTOME pathways (https://reactome.org/) in IES172 and IES68 signature genes (*clusterProfiler* package in R), which were ranked based on the gene ratio (gene count divided by set size). (B) Dot plot showing the average expression of IES genes (RNA-sequencing signature [IES172]; NanoString IES signature [IES68], LASSO-based RNA-sequencing IES signature [PI24] and LASSO-based NanoString IES signature [PI20]) on immune cell types identified in 8 single-cell RNA-sequencing AML samples. Data were retrieved from the Synapse data repository (https://www.synapse.org/#!Synapse:syn21991338; SynapseID: syn21991338) (Dufva et al., 2020) and analyzed with R v.4.0.2. MPP = multipotent progenitors; MEP = megakaryocyte erythroid progenitors; GMP = granulocyte-macrophage progenitors; CMP = common myeloid progenitors; NK = natural killer. (C) Correlation between the IES172 score and previously published immune traits (n=45) (Danaher et al., 2017; Vadakekolathu et al., 2020b). Signature scores are available through the original publications. (D) Correlation between IES68 scores and leukemia burden at diagnosis in the PMCC cohort. Data were compared using the Mann Whitney *U* test for unpaired determinations. BM = bone marrow; PB = peripheral blood. (E) Response to induction chemotherapy in patients with above median and below median prognostic index (PI20). PIF = primary induction failure following a standard 1 or 2 cycles of induction chemotherapy. CR = complete remission (defined as <5% BM blasts). (F) Kaplan-Meier estimates of relapse-free survival (RFS) in PMCC patients with above median (magenta line) and below median (blue line) PI. Survival curves were compared using a log-rank test (*survminer* package in R). HR = hazard ratio. (G) Kaplan-Meier estimates of OS in PMCC patients with higher than median (magenta line) and lower than median (blue line) PI. Survival curves were compared using a log-rank test (*survminer* package in R). (H) AUROC curve measuring the predictive ability of the 20 IES genes (blue line) and the ELN cytogenetic risk classifier (magenta line) for overall survival (OS). AUROC = 1.0 denotes perfect prediction and AUROC = 0.5 denotes no predictive ability. (I) Cox proportional hazards (PH) model for OS in the PMCC AML cohort. PI20 = prognostic index; LSC17 = leukemia stem cell 17 score; B = beta coefficient; SE = standard error; DF = degrees of freedom; Sig. = significance; Exp(B) = odds ratio.

Using LASSO penalized regression for feature selection and collinearity reduction, we identified 20 genes in the NanoString IES68 signature that were predictive of OS. We then computed a PI20 using expression values for these genes and β coefficients previously derived from Cox proportional hazards (PH) models of the TCGA-AML discovery cohort. The 20-gene PI (PI20) was associated with primary induction failure (PIF) in response to standard chemotherapy (**Figure 4E**). A high PI20 was associated with significantly shorter RFS and OS in the Princess Margaret Cancer Center (PMCC) cohort (*P*<0.001 for both; **Figure 4F-G**). Overall, the 20-gene PI predicted OS with greater accuracy (AUROC value of 0.847) than the 2017-ELN cytogenetic risk classifier (AUROC value of 0.643; **Figure 4H**). These observations were validated in an independent AML series including subjects with PIF enrolled in the AMLCG-2008 study (GSE106291; n=250 patients; **Figure S9**) (Herold et al., 2018).

In contrast to the recently defined IFN-γ gene signature (Vadakekolathu et al., 2020c), the 20-gene PI score separated survival within each cytogenetically-defined risk group (**Figure S10A**), as well as after censoring at time of hematopoietic stem cell transplantation (**Figure S10B**). The latter finding suggests that differences in clinical outcomes between PI20^high^ and PI20^low^ cases were not merely attributable to treatment intensity.

We calculated the LSC17 score for the PMCC cohort using publicly available gene expression data (GSE76004) and the same weights as those provided in the original publication (**Figure S11A-B**) (Ng et al., 2016). In line with TCGA data, the LSC17 score separated RFS and OS in both PI20^low^ (**Figure S11C-S11D**) and PI20^high^ cases (**Figure S11E-S11F**). Specifically, patients with high 20-gene PI, who experienced a 5-year OS rate of 11% (**Figure 4G**), were further dichotomized into a subgroup of LSC17^low^ individuals with an improved 5-year OS probability of 55% (**Figure S11F**). Furthermore, when stratifying patients in the LSC17^high^ and LSC17^low^ subgroup by the PI20, we identified a subset of LSC17^high^ subjects with very high-risk AML, who had 5-year RFS and OS rates of only 10% and 3.5%, respectively (**Figure S11G-S11J**).

We formally tested the interaction between senescence- and stemness-related pathways by a multiplication term in the Cox PH model. As shown by the Wald χ^2^ statistics (**Figure 4I**), the 20-gene PI was substantially more predictive of OS (*P*<0.001) in this modeling framework than the LSC17 score (*P*=0.001). In addition, the interaction between the two continuous variables was statistically significant (*P*=0.013), indicating that a higher PI20 will increase the association between the LSC17 score and OS. Taken together, these analyses suggest that the 20-gene PI and its integration with the LSC17 score could provide accurate prognostic risk stratification.

### IES scores predict survival in independent pediatric AML cohorts

Microenvironmental immune gene sets are known to be differentially expressed between children and adults with AML (Vadakekolathu et al., 2020b; Willier et al., 2021), which may in part be due to differences in pediatric versus adult AML biology (Bolouri et al., 2018; Conneely and Stevens, 2021; de Rooij et al., 2015; Tarlock and Meshinchi, 2015). Furthermore, immunosenescence, a process of remodeling of immune functions upon chronic antigen exposure, is associated with physiologic aging (Carrasco et al., 2021; Mittelbrunn and Kroemer, 2021). We thus examined the relevance and applicability of the IES score to childhood AML and first analyzed diagnostic BM samples from 145 pediatric patients with *de novo* AML in the Children’s Oncology Group (COG)-TARGET AML cohort for whom RNA-sequencing data are publicly available (Bolouri et al., 2018). The IES172 score correlated inversely with leukemia burden (**Figure 5A-5B**) and was significantly higher at time of relapse (n=31 paired BM samples; **Figure 5C**). Importantly, an above median 24- gene PI predicted significantly worse RFS (*P*=0.0044) and OS (*P*=0.018; **Figure 5D-5E**). We then retrieved NanoString transcriptomic data from an additional cohort of pediatric subjects with AML (CHOP series, n=40 patients; GSE134589) (Vadakekolathu et al., 2020b). In line with previous results in adult AML (**Figure 2D**), the IES68 score was higher in children with an immune infiltrated/activated AML (**Figure 5F**) and in BM samples obtained at time of response assessment compared with disease onset (**Figure 5G**). Finally, the 20-gene PI separated patients into subgroups with different RFS and OS probabilities (**Figure 5H-5I**). These data support the applicability of IES scores in childhood AML as well.

**Figure 5.**
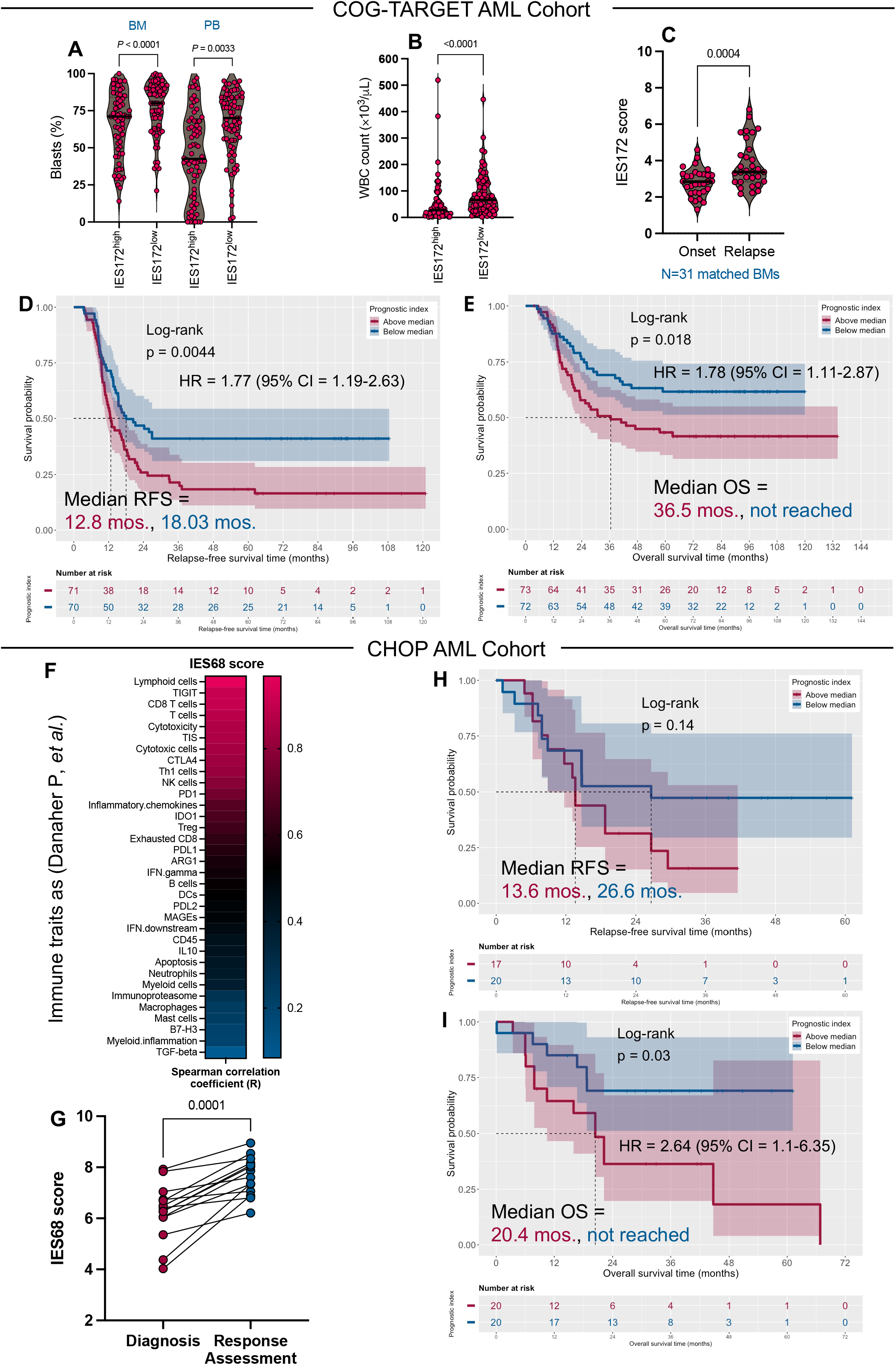
Immune effector senescence (IES) scores correlate with immune infiltration and separate survival in pediatric AML cohorts. (A) Leukemia burden in COG-TARGET AML cases (n = 145) with above median and below median IES172 scores. Data were compared using the Mann Whitney *U* test for unpaired determinations. BM = bone marrow; PB = peripheral blood. (B) White blood cell (WBC) count at diagnosis in COG-TARGET AML cases with above median and below median IES172 scores. Data were compared using the Mann Whitney *U* test for unpaired determinations. (C) IES172 scores at time of AML diagnosis and response assessment (gene expression data from matched BM samples available in 31 COG-TARGET AML cases). (D) Kaplan-Meier estimate of relapse-free survival (RFS) in patients from the COG-TARGET AML cohort with above median (magenta line) and below median (blue line) prognostic index (PI24). Survival curves were compared using a log-rank test (*survminer* package in R). HR = hazard ratio. (E) Kaplan-Meier estimate of overall survival (OS) in patients from the COG-TARGET AML cohort with above median (magenta line) and below median (blue line) PI24. (F) Correlation between the IES68 score and previously published immune traits (n=45) in the CHOP AML series. Signature scores are available through the original publications (Danaher et al., 2017; Vadakekolathu et al., 2020b). (G) IES68 scores in samples from the CHOP AML series collected at time of diagnosis and response assessment (n = 14 matched BM samples). Data were compared using the Wilcoxon matched pairs signed rank test. (H) Kaplan-Meier estimates of RFS in patients from the CHOP AML cohort (n = 40) with above median (magenta line) and below median (blue line) PI20. (I) Kaplan-Meier estimate of OS in patients from the CHOP cohort with above median (magenta line) and below median (blue line) PI20. HR = hazard ratio.

### IES scores are increased at time of response assessment

To examine further the impact of induction chemotherapy on IES scores, we generated nCounter gene expression data using serial BM samples from a large cohort of patients with newly diagnosed AML (SAL and JHU2, totaling 90 patients and 183 BM specimens longitudinally collected at time of diagnosis and response assessment) (**Table S1**). The IES68 scores were significantly higher after chemotherapy (**Figure 6A-6B**). As expected, the 20-gene PI separated both RFS (*P*=0.011; **Figure 6C**) and OS (*P*=0.0015; **Figure 6D**) in this cohort.

**Figure 6.**
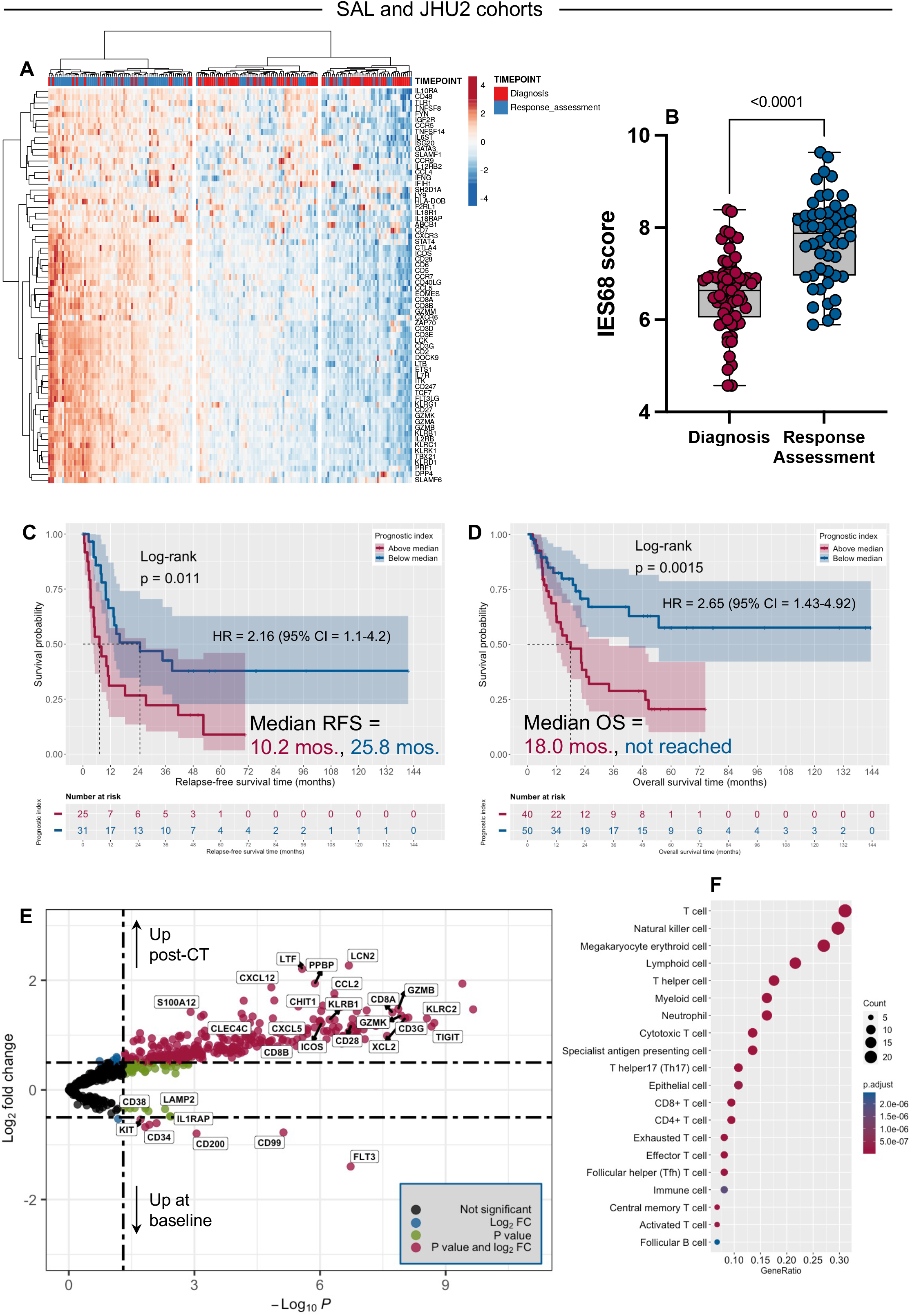
Immune effector senescence (IES) scores increase at time of response assessment and predict outcomes in additional external AML cohorts. (A) Expression of the IES68 genes in patients from the SAL and JHU cohorts (n = 183 BM samples from 90 patients). ClustVis, an online tool for clustering of multivariate data (Euclidean distance, complete linkage), was used for data visualization (Metsalu and Vilo, 2015). The heatmap annotation track shows sample collection timepoints (baseline and post-chemotherapy response assessment). (B) IES68 score at time of diagnosis and response assessment. Data were compared using the Mann-Whitney *U* test for unpaired determinations. (C) Volcano plot showing differentially expressed genes (DEGs) between samples collected at baseline and post-chemotherapy (CT) response assessment (*EnhancedVolcano* package in R). Genes discussed in the paper are highlighted using black boxes. (D) Graphical summary of over-representation analysis (ORA; *clusterProfiler* package in R) showing the overlap between DEGs (post-chemotherapy *versus* baseline) and curated cell type signature gene sets (C8 collection), which were retrieved from the MSigDB (http://www.gsea-msigdb.org/gsea/index.jsp). Gene ratio = gene count divided by set size. (E) Kaplan-Meier estimate of relapse-free survival (RFS; data available in 56 subjects) in higher than median (magenta line) and lower than median (blue line) PI20 groups. Survival curves were compared using a log-rank test (*survminer* package in R). HR = hazard ratio. (F) Kaplan-Meier estimate of overall survival (OS; data available in 90 subjects) in higher than median (magenta line) and lower than median (blue line) PI20 groups. Survival curves were compared using a log-rank test (*survminer* package in R).

Differential expression analysis revealed upregulation of T-cell/NK-cell genes (*CD3G*, *CD8A*, *CD8B*, *CD28*, *GZMK*, *GZMB*), co-signaling molecules (*KLRC2*, *KLRB1*, *TIGIT*, *CD40L*, *ICOS*), genes involved in myeloid (*LCN2*, *LTF*, *S100A12*) and dendritic cell (*CHIT1*, *CLEC4C*) differentiation and in chemoattraction (*CXCL5*, *CXCL12*, *CCL2*, *PPBP*, *XCL2*) after chemotherapy (**Figure 6E**). As expected, genes associated with AML proliferation (*FLT3*, *KIT*), leukemia stem cells (*CD34*, *CD38*, *IL1RAP*), and candidate genes overexpressed in AML (*CD99*, *CD200* and *LAMP2*) were downregulated after chemotherapy, consistent with recent data using scRNA-seq and IHC after induction chemotherapy (van Galen et al., 2019). GSEA on the C2, C7 and C8 gene sets from the MSigDB revealed over-representation of T-cell subsets, NK-cell and antigen presenting cell signatures (**Figure 6F**) and clearance of leukemia signatures after chemotherapy (**Figure S12A**). Furthermore, oncogenic pathways were downregulated, while immune signatures and pathways were enriched at time of response assessment (**Figure S12B**).

### IES genes define ICB-unresponsive TMEs both in AML and in solid tumors

We assessed the relevance of IES scores in relation to therapeutic response to ICB. We profiled primary BM samples from 33 adult patients with newly diagnosed or relapsed/refractory AML who were treated with AZA+Pembro (clinicaltrials.gov identifier: NCT02845297; **Table S5**; GSE178926). We examined DEGs at baseline between patients who subsequently achieved CR and those who were non-responders. Using unsupervised hierarchical clustering of DEGs (**Figure 7A**), two patient clusters were observed. Cluster 1 (C1 in **Figure 7A**) was enriched for patients who achieved CR (∼63%) and for patients with PI20 scores below median (∼63%). In contrast, only ∼14% of patients in Cluster 2 (C2 in **Figure 7A**) achieved CR and ∼21% of them had below-median PI20. Notably, patients with low PI20 experienced prolonged OS (median = 15.6 months compared with 4.1 months in patients with high PI20; *P*=0.011; **Figure 7B**). We also observed heightened expression of type I and type II IFN signaling genes (*IRF8*, *IFNA1*, *IFNA17*, *CXCL10*, *CCL20*) in the PI^low^ group (**Figure 7A**), prompting us to examine the ability of a published IFN signature to predict OS in patients treated with AZA+Pembro (Vadakekolathu et al., 2020b). As shown in **Figure 7C**, high IFN scores were associated with prolonged OS (*P*=0.01). Taken together, these data reveal the unique ability of IES genes to define both chemotherapy- and ICB- unresponsive AML TMEs. By contrast, IFN-γ–related genes have been previously shown to be associated with chemotherapy resistance while also predicting response to T-cell engagers (Uy et al., 2021; Vadakekolathu et al., 2020b).

**Figure 7.**
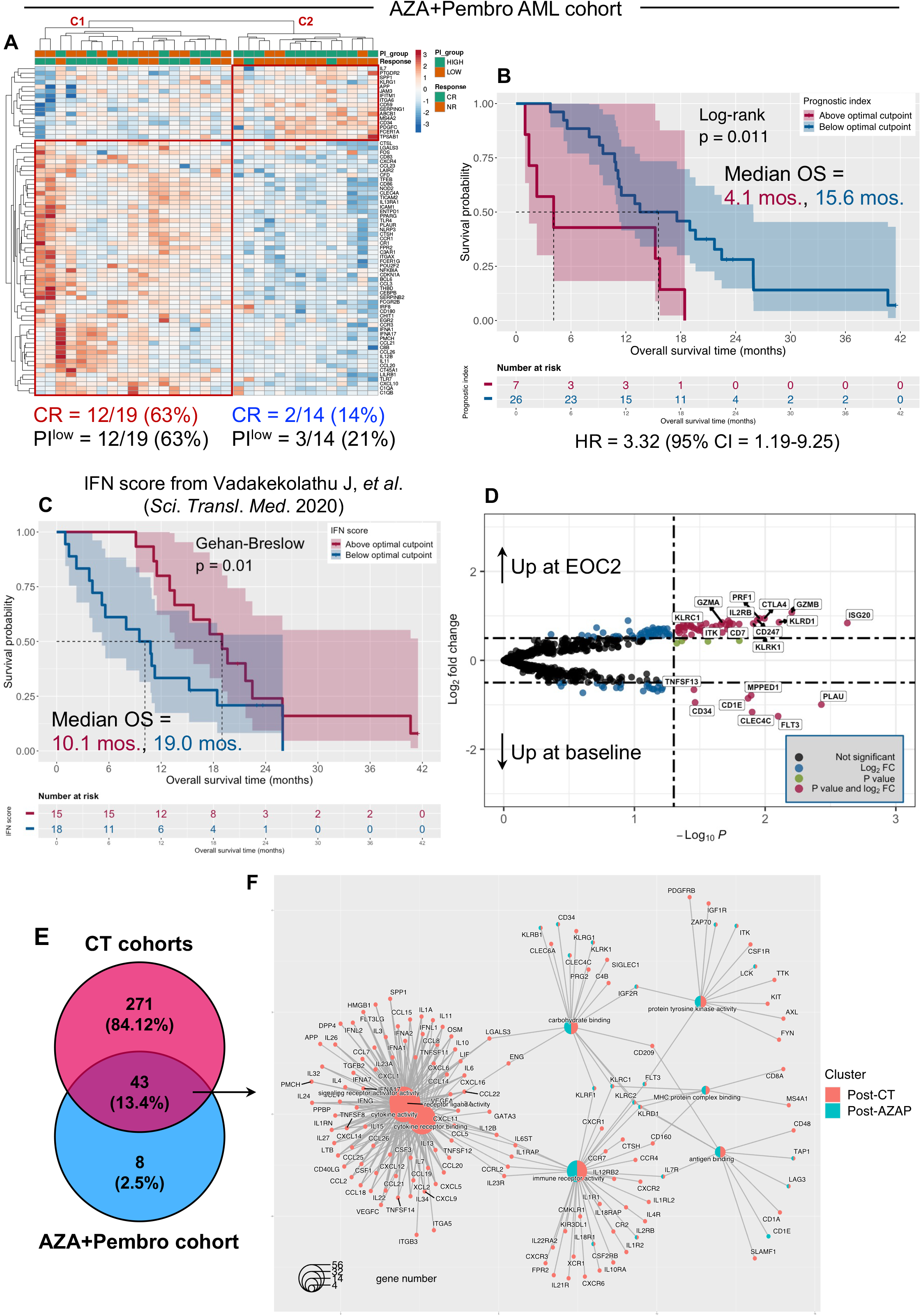
Immune effector senescence (IES) scores predict response to AZA+Pembro in clinical trial NCT02845297. (A) Differentially expressed (DE) genes at baseline associated with complete response (CR) to AZA+Pembro. ClustVis, an online tool for clustering of multivariate data (Euclidean distance, complete linkage), was used for data visualization (Metsalu and Vilo, 2015). The heatmap annotation track shows the prognostic index (PI20) group and response status (complete remission [CR] and non-responder [NR]) after 2 cycles of azacitidine and pembrolizumab. Complete response was defined as CR, CR with partial hematologic recovery (CRh), CR with incomplete hematologic recovery (CRi), or morphological leukemia-free state (MLFS) at the end of cycle 2. Patients with partial response (PR; >50% decrease in bone marrow (BM) blasts from baseline to 5% to 25% at the end of cycle 1) were categorized as NRs. (B) Kaplan-Meier estimate of overall survival (OS) in patients with above median (magenta line) and below median (blue line) PI20. Survival curves were compared using a log-rank test (*survminer* package in R). HR = hazard ratio. (C) Kaplan-Meier estimate of overall survival (OS) in patients with above median (magenta line) and below median (blue line) IFN scores, which were computed as previously published (Vadakekolathu et al., 2020b). Survival curves were compared using the Gehan-Breslow-Wilcoxon test (*survminer* package in R), a generalization of the Wilcoxon rank-sum test which gives more weight to deaths at early time points. (D) Volcano plot showing differentially expressed genes (DEGs) between baseline and end-of-cycle 2 (EO2) BM samples from patients in the AZA+Pembro cohort (*EnhancedVolcano* package in R). The top 20 DEGs are shown in black boxes. (E) Venn diagram showing the overlap between DEGs post-treatment *versus* baseline in the chemotherapy (CT; SAL and JHU) and AZA+Pembro cohorts. (F) Non-redundant, enriched gene ontologies in DEGs between the chemotherapy (CT) and AZA+Pembro cohorts were visualized as a network diagram (cnetplot) with color nodes using the cnetplot function of the *GOSemSim* package in R (Yu et al., 2010).

We sought to identify genes at the intersection of responses to chemotherapy and AZA+Pembro. We examined DEGs between matched post-treatment (available in 31 patients after cycle 2) *versus* pre-treatment BM samples in the immunotherapy cohort. Treatment with AZA+Pembro resulted in upregulation of immune effector-related genes (*GZMA*, *GZMB*, *PRF1*, *KLRD1*, *NCR1*), T-cell and NK co-signaling molecules (*CTLA4*, *KLRB1*, *KLRC1*, *KLRC2*, *KLRK1*), cytokine receptors (*IL7R*, *IL2RB*), IFN responsiveness (*ISG20*) and T-cell signaling genes (*CD274*, *ITK*, *CD7*, *ZAP70*) (**Figure 7D**). As with the chemotherapy cohort (**Figure 6E**), AZA+Pembro treatment was associated with downregulation of leukemia-associated genes (*FLT3*, *CD34*). We identified 43 genes that were significantly differentially expressed in both post-chemotherapy and post-AP BM samples (**Figure 7E**). We assessed the semantic distance between GOs corresponding to these 43 genes using the GOSemSim Bioconductor R package (Yu et al., 2010). This procedure, which measures GO and gene similarity thereby minimizing the redundancy of GO categorization, identified shared nodes, which included GO terms linked to immune functions, as well as a prominent “macro-cluster” which was unique to the chemotherapy setting. This macro-cluster encompassed GO terms and genes related to cytokine activity and cytokine receptor signaling (**Figure 7F**).

To investigate how generalizable the above findings are to solid tumors, we conducted an exploratory analysis of IES and its correlation with response to ICB in melanoma. We calculated the 24-gene PI for patients in the TCGA Pan-Cancer Atlas profiling project (441 subjects with resected primary and/or metastatic melanoma who received no previous systemic therapy) (Cancer Genome Atlas Network, 2015). The PI24 was not correlated with patient age or tumor mutation count (**Figure S13A-S13B**) and was lower in patients with an immune-enriched TME and with high expression of immune-associated functional gene signatures (Bagaev et al., 2021) (**Figure S13C-S13D**). Interestingly, the PI24 refined the ability of the immune-enriched, ICB-responsive TME profile, but not the depleted TME subtype (Bagaev et al., 2021), to stratify patient survival (**Figure S13E-S13F**). As observed above in AML, PFS and OS rates were lower for melanoma cases with high PI24 (**Figure 8A-8B**).

**Figure 8.**
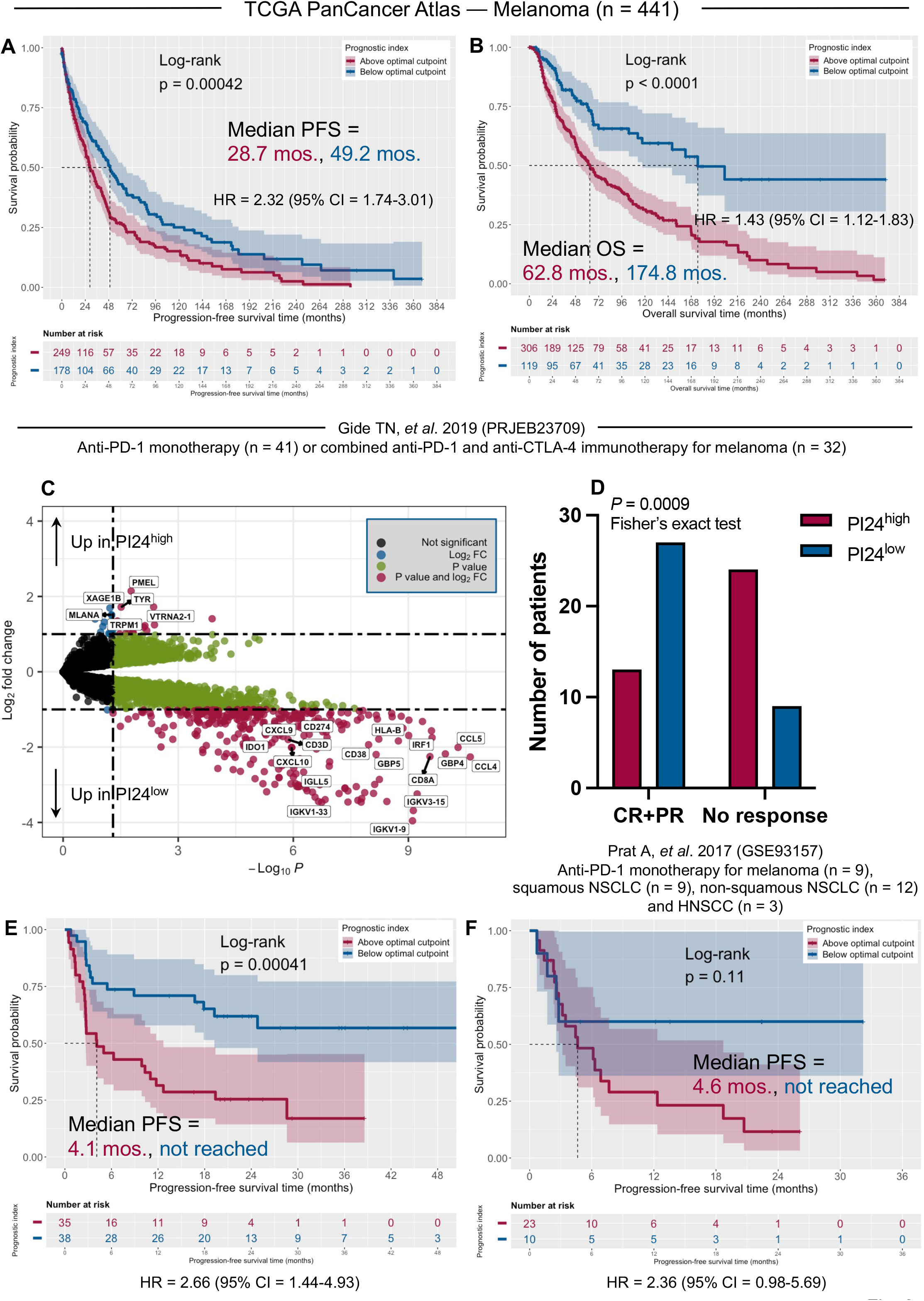
Immune effector senescence (IES) scores predict immunotherapy response in melanoma. (A) Progression-free survival (PFS) in 441 patients with melanoma from the TCGA Pan-Cancer Atlas profiling project. Subjects were stratified based on an optimal cut-point of the PI24, which was computed using the *maxstat* package in R. Survival curves were compared using a log-rank test (*survminer* package in R). RNA-sequencing and outcomes data were retrieved through the cBioPortal for Cancer Genomics (https://www.cbioportal.org/). HR = hazard ratio. (B) Overall survival (OS) in patients with melanoma from the TCGA Pan-Cancer Atlas cohort. (C) Volcano plot (*EnhancedVolcano* package in R) showing differentially expressed genes (DEGs) between PI24^high^ and PI24^low^ patients in the PRJEB23709 immunotherapy cohort [73 subjects with melanoma treated with standard-of-care single-agent nivolumab or pembrolizumab (n = 41) or combination anti-PD-1 + anti-CTLA-4 (n = 32; Table S9)]. RNA-sequencing and outcomes data were retrieved through the original publication (Gide et al., 2019) and the Tumor Immune Dysfunction and Exclusion (TIDE) portal (http://tide.dfci.harvard.edu/login/) (Jiang et al., 2018). The top 15 DE genes are shown in black boxes. (D) Number of responders and non-responders with above median and below median PI24 in the PRJEB23709 immunotherapy cohort. Fisher’s exact test. CR = complete response; PR = partial response. In the original publication (Gide et al., 2019), responders are defined as individuals with complete response, partial response, or stable disease of greater than 6 months with no progression, whereas non-responders are defined as progressive disease or stable disease for less than or equal to 6 months before disease progression. (E) PFS in patients with melanoma in the PRJEB23709 immunotherapy cohort. Patients were dichotomized based on an optimal cut-point of PI24 values. (F) PFS in patients with advanced solid tumors (n = 9 with melanoma, n = 9 with squamous non-small cell lung carcinoma [NSCLC], n = 12 with non-squamous NSCLC, n = 3 with head and neck squamous cell carcinoma [HNSCC]) treated with anti-PD-1 monotherapy in the GSE93157 series. Patients were dichotomized based on an optimal cut-point of PI20 values. NanoString profiling data and outcomes data were retrieved through the original publication (Prat et al., 2017) and the TIDE portal (Jiang et al., 2018).

Finally, we analyzed publicly available RNA-sequencing data from 73 melanoma patients treated with standard-of-care single-agent nivolumab or pembrolizumab (n = 41) or combination anti-PD-1 + anti-CTLA-4 (n=32; PRJEB23709; **Table S6**) (Gide et al., 2019). In this series, patients with above-median PI24 showed enrichment in melanocyte-associated markers (*MLANA*, *TYR*, *PMEL*; **Figure 8C**) and poor response to ICB based on RECIST criteria (**Figure 8D**). As with TCGA Pan-Cancer Atlas data, samples from patients with below-median PI24 expressed high levels of immunoglobulin genes, *CD8A*, and chemokine genes (*CCL4*, *CCL5*, *CXCL10*; **Figure 8C**), and correlated with significantly higher PFS rates (*P*=0.00041; **Figure 8E**). Similar results were observed in an independent immunotherapy cohort (GSE93157; **Table S7**) comprising 33 patients with melanoma, head and neck squamous cell carcinoma or lung cancer (Prat et al., 2017), where individuals with low and high PI20 experienced PFS rates of 60% and 12%, respectively, in response to anti-PD-1 monotherapy (**Figure 8F**). Overall, these findings suggest that signatures of IES might also be applied as potential biomarkers of response to ICB in solid tumors.

## Discussion

An unanswered question in AML is whether deranged T-cell functions affect the likelihood of therapeutic response to chemotherapy and/or immunotherapy. Our prior efforts to characterize the AML immune TME using transcriptomic and spatial profiling approaches led to the discovery of an IFN-γ-dominant and inflamed BM milieu (Rutella et al., 2018; Uy et al., 2021; Vadakekolathu et al., 2020b). In the present study, features of immune effector senescence were identified in multiple independent cohorts of adult and pediatric patients with AML and were found to be associated with leukemia stemness and with poor response to induction chemotherapy. The derived IES gene set improved OS prediction afforded by clinically validated cytogenetic categories and by the experimental LSC17 signature (Dohner et al., 2017; Ng et al., 2016) and defined ICB-unresponsive microenvironments.

Determining how dysfunctional T-cell states modulate therapeutic response or resistance in AML remains a challenge, partly due to lack of selective markers that parse exhaustion from senescence (Akbar and Henson, 2011; Liu et al., 2020). We previously detected increased numbers of circulating senescent-like T cells in AML, which were associated with low likelihood of response to induction chemotherapy (Knaus et al., 2018).

Some reports suggest that tumors induce T-cell senescence *via* cancer cell-derived soluble molecules while others implicate CD4^+^ regulatory T cells in this process (Liu et al., 2018; Montes et al., 2008). Herein, we found that AML blasts influence T-cell activation and proliferation through direct contact and through bystander modulation, whereas induction of CD8^+^ T-cell senescence appears primarily dependent on the latter. These mechanisms are particularly relevant for hematologic malignancies such as AML, since leukemia blasts are proximate to circulating T cells and, as such, their potential to promote T-cell senescence is expected to be greater than peripherally located solid tumors.

It has also been shown that chemotherapy-induced senescence confers higher tumor-initiating potential to AML and solid tumor cell lines *versus* non-senescent tumor cells (Duy et al., 2021; Milanovic et al., 2018). While we observed an association between stemness and effector senescence programs, an important question to be addressed is whether crosstalk between senescent-like AML cells and immune effectors could potentially amplify immunosuppressive circuits leading to failed control of residual disease. Senescent-like cells are known to secrete inflammatory chemokine, cytokines, and growth factors in a paracrine fashion, promoting the reprogramming of neighboring cells (Abdul-Aziz et al., 2019; Mosteiro et al., 2016; Ritschka et al., 2017). Furthermore, the humoral communication *via* senescence-associated secretory phenotype factors might accelerate tumor progression by maintaining chronic inflammation (Lee and Schmitt, 2019). In accord with this model, we show that IES signatures that are shared between CD8^+^ T cells and NK cells increase after chemotherapy. In contrast to T-cell exhaustion, IES states are maintained by intrinsic signaling induced by DNA damage or other stress responses (Akbar et al., 2016; Liu et al., 2018; Mondal et al., 2013). While a subset of the IES signature comprised exhaustion genes, the overlap between the IES score and published T-cell exhaustion gene sets was minimal (Guo et al., 2018; Miller et al., 2019). Enhancing T cell-mediated clearance of AML is an attractive therapeutic strategy, but immunotherapy trials in AML using BiTE constructs and some trials with ICB have met with only limited success (Ravandi et al., 2020; Zeidan et al., 2021; Zeidan et al., 2018). Multiple mechanisms have been proposed to explain AML resistance to therapeutic attempts to reverse T-cell exhaustion by ICB. These include upregulation of alternative checkpoint receptors or diminished T-cell infiltration in patients with advanced disease (Daver et al., 2021; Vago and Gojo, 2020). Our data suggest that senescent-like T cells in pre-treatment BM samples are unable to lyse AML blasts when activated with the CD3/CD33 BiTE construct. Consistent with this, a higher proportion of senescent-like CD8^+^ T cells in the BM and blood was associated with lower response rates to ICB with pembrolizumab sequenced after high-dose cytarabine in relapsed/refractory AML (Zeidner et al., 2021). Therefore, this T-cell population may underpin resistance to immunotherapy.

Our study also shows that the initially defined PI in newly diagnosed AML also predicts outcomes in patients receiving AP immunotherapy, and encourages the pursuit of senescence reversal as a strategy to functionally reinvigorate T cells and to improve response rates to ICB and other T cell-targeting immunotherapies (Uy et al., 2021; Zeidner et al., 2021). Strategies to reverse senescence with senolytics are currently being tested in animal models (Lee S, et al. 2021). By analyzing the immune transcriptome of pre-treatment samples from the AZA+Pembro cohort, we identify gene sets and biological functions enriched in responders. In contrast to the IES score, the IFN-γ signature score was associated with response to ICB. A plausible explanation for this observation is that stemness states negatively affect type I IFN signaling and anti-cancer immunity, ultimately leading to poor AML-cell killing (Miranda et al., 2019). The IES-related gene set also predicted long-term outcomes and objective responses to single-agent nivolumab or pembrolizumab, or to combination anti-PD-1 + anti-CTLA-4 in melanoma (a tumor type known to derive durable clinical benefit from ICB) (Fairfax et al., 2020; Gide et al., 2019; Huang et al., 2019; Prat et al., 2017). Future prospective immunotherapy clinical trials are warranted to validate the translational relevance of the IES signature.

One limitation of our study is that we focused primarily on gene sets pertaining to immune biology. However, efforts to link immunology with genomic subtypes, therapeutic response and clinical outcomes in AML are in their infancy (Bruck et al., 2020; Dohner et al., 2021; Dufva et al., 2020; Vadakekolathu et al., 2020a; Vadakekolathu et al., 2020b). In contrast, genome-wide transcriptomic approaches and high-dimensional single-cell analyses have been extensively employed to resolve the molecular heterogeneity and clonal diversity of malignant AML cells (Horibata et al., 2019; Miles et al., 2020; Morita et al., 2020; Toffalori et al., 2019; van Galen et al., 2019). scDNA-seq and scRNA-seq studies would be required to explore relations between T-cell differentiation stages, clonal complexity and AML hierarchies (Miles et al., 2020; van Galen et al., 2019), but a major challenge is the difficulty of acquiring adequate numbers of T cells from TMEs in which cells of the myeloid lineage are predominant.

Overall, our findings offer advantages over signatures of T-cell exhaustion, which are solely predictive of response to ICB (Im et al., 2016; Jiang et al., 2018; Sade-Feldman et al., 2018). Our approach elucidates the immune contexture of AML in both chemotherapy and ICB settings, enables refinement of risk stratification and generates hypotheses for further investigation and clinical exploration of strategies to overcome immunosenescence.

## Supplemental information

Supplemental information can be found online.

## Data Availability

The transcriptomic datasets generated in this study have been deposited on to the GEO repository under accession numbers GSE176100 and GSE178926 and will be publicly available as of the date of publication.

https://www.ncbi.nlm.nih.gov/gds

## STAR Methods

Detailed methods are provided in the online version of this paper and include the following:

- Key Resources Table
- Resource Availability

- Lead Contacts
- Materials Availability
- Data and Code Availability
- Subject Details

- Wet-lab cohorts
- Immunotherapy cohort
- Data sources for *in silico* analyses (AML)
- Data sources for *in silico* analyses (melanoma)
- Method Details

- *In vitro* cytotoxicity assays
- RNA isolation and processing
- nCounter data quality control and normalization
- Signature calculation
- GSEA and leading-edge analysis
- Single-cell RNA sequencing data analysis
- Statistical Analysis
- Additional References

## Acknowledgements

S.R., J.V. and S. Reeder were supported for these studies by the Qatar National Research Fund (NPRP8-2297-3-494), the John and Lucille van Geest Foundation and Nottingham Trent University’s School of Science and Technology. S.K.T. was supported for these studies by NIH/NCI 1U01CA232486 and U01CA243072, Department of Defense Translational Team Science Award CA180683P1, Andrew McDonough B+ Foundation, Gabrielle’s Angel Foundation for Cancer Research, Rally Foundation for Childhood Cancer Research, and the St Baldrick’s Foundation/Stand Up to Cancer Pediatric Dream Team. Stand Up to Cancer is a program of the Entertainment Industry Foundation administered by the American Association for Cancer Research. Childhood leukemia specimen banking was supported by the Children’s Hospital of Philadelphia’s Center for Childhood Cancer Research. L.L. was supported for these studies by NIH/NCI P01CA225618.

## Author contributions

Concept and design: S. Rutella, L.L.

Development of methodology: S. Rutella, J.V., F.M., T.Y., R.M., S. Reeder, V.R., L.L.

Acquired, consented, and managed patients; processed patient samples: R.M., H.A., M.K., H.K., J.F.Z., A.A., M.D.M., S.K.T., M.B., I.G., L.L.

Analysis and interpretation of data: S. Rutella, J.V., F.M., S. Reeder, T.Y., R.M., B.D., B.R.B., V.R., M.D.M., S.K.T., M.B., I.G., L.L.

Writing of the manuscript: S. Rutella

Review and/or revision of the manuscript: S.R., J.V., F.M., R.M., B.D., B.R.B., J.F.Z., S.K.T., M.B., I.G., L.L.

Study supervision: S. Rutella, L.L.

## Declaration of interest

I.G. received research support from Merck (clinical study #NCT02845297).

L.L. received research support from Genentech (imCORE grant #ML40354).

## Notes

### Author Declarations

Primary patient specimens (non-promyelocytic AML) and associated clinical data were obtained via informed consent in accordance with the Declaration of Helsinki on research protocols approved by the Ethics Committee of TU Dresden and Studienallianz Leukaemie (Germany) (EK98032010) and by the Childrens Hospital of Philadelphia (10-007767) and Johns Hopkins University (JHU) Institutional Review Boards.

