## Supplemental methods for "Signatures of immune senescence predict outcomes and define checkpoint blockade-unresponsive microenvironments in acute myeloid leukemia"

**STAR METHODS**

**KEY RESOURCES TABLE**

| **Reagent or Resource** | **Source** | **Identifier** |
| --- | --- | --- |
| Antibodies |  |  |
| CD45 (clone 2D1) | BioLegend | Cat#: 368510 |
| CD19 (clone HIB19) | BioLegend | Cat#: 302258 |
| CD4 (clone OKT4) | BioLegend | Cat#: 317408 |
| CD8 (clone SK1) | BioLegend | Cat#: 344740 |
| CD8 (clone RPA T8) | e-Bioscience | Cat#: 45-0088-42 |
| CD57 (clone HNK1) | BioLegend | Cat#: 359608 |
| KLRG1 (clone SA231A2) | BioLegend | Cat#: 367716 |
| CD28 (clone CD28.2) | BioLegend | Cat#: 302946 |
| CD33 (clone P67.6) | BioLegend | Cat#: 366622 |
| TIGIT (clone A15153G) | BioLegend | Cat#: 372712 |
| CD127 (clone A019D5) | BioLegend | Cat#: 351326 |
| PD1 (clone EH12.1) | BD Biosciences | Cat#: 560795 |
| Tim-3 (clone F38-2E2) | BioLegend | Cat#: 345034 |
| CD34 (clone 561) | BioLegend | Cat#: 343608 |
| CD3 (clone OKT3) | BioLegend | Cat#: 317340 |
| Ki-67(clone B56) | BD Biosciences | Cat#: 561284 |
| γH2AX (clone N1-431) | BD Biosciences | Cat#: 562377 |
| ICOS (clone C398.4) | BioLegend | Cat#: 313534 |
| CD25 (clone BC96) | e-Bioscience | Cat#: 25-0259-42 |
| Biological Samples |  |  |
| Newly diagnosed AML | This paper | GEO: GSE176100 |
| Relapsed/refractory AML | This paper | GEO: GSE178926 |
| Newly diagnosed AML | This paper | Flow cytometry |
| Deposited data |  |  |
| Newly diagnosed AML | Vadakekolathu et al. (2020) | GEO: GSE134589 |
| Newly diagnosed AML | TCGA-AML | https://www.cbioportal.org/  See **Table S1** |
| Newly diagnosed AML | COG-TARGET AML | https://www.cbioportal.org/  See **Table S1** |
| Newly diagnosed AML | Beat-AML Master Trial | https://www.cbioportal.org/  See **Table S1** |
| Newly diagnosed AML | Dufva et al. (2020) | syn21991338 |
| Newly diagnosed AML | Herold et al. (2018)  Li et al. (2013) | GEO: GSE37642 |
| Newly diagnosed AML | Ng et al. (2016) | GEO: GSE76004 |
| Newly diagnosed AML | Herold et al. (2018) | GSE106291 |
| Cutaneous melanoma | TCGA PanCancer Atlas  Gao et al. (2013) | https://www.cbioportal.org/ |
| Cutaneous melanoma immunotherapy | Gide et al. (2019) | ENA: PRJEB23709  See Table S9 |
| Cutaneous melanoma immunotherapy | Prat et al. (2017) | GEO: GSE93157  See Table S10 |
| Cutaneous melanoma | Bagaev et al. (2021) | DOI: 10.1016/j.ccell.2021.04.014  https://science.bostongene.com/tumor-portrait/ |
| Chemicals, Peptides, and Recombinant Proteins |  |  |
| IL3 | PHC0034 | Life Technologies |
| G-CSF | RGCSF10 | Life Technologies |
| GM-CSF | PHC2013 | Life Technologies |
| SCF | PHC2115 | Life Technologies |
| Critical Commercial Assays |  |  |
| RNA extraction kit | Qiagen | Cat#: 74106 |
| PanCancer Immune Profiling kit | NanoString Technologies | Cat#: 115000132 |
| Qubit™ RNA HS Assay Kit | Thermo Fisher Scientific | Cat#: Q32852 |
| RNA Clean & Concentrator-5 with DNase I Set | Zymo Research | Cat#: R1013 |
| Software and Algorithms |  |  |
| CIBERSORT | Gentles et al. (2015) | https://cibersort.stanford.edu/ |
| clusterProfiler R package | Yu et al. (2012) | https://guangchuangyu.github.io/software/clusterProfiler/ |
| ClustVis | Metsalu et al. (2015) | https://biit.cs.ut.ee/clustvis/ |
| corrplot R package | Wei et al. (2021) | https://github.com/taiyun/corrplot |
| EnhancedVolcano R package | Blighe et al. (2019) | DOI: 10.18129/B9.bioc.EnhancedVolcano |
| EPIC | Racle et al. (2017) | http://epic.gfellerlab.org |
| GeneMANIA | Warde-Farley et al. (2010) | https://genemania.org/ |
| GEPIA2021 | Chenwei et al. (2021) | http://gepia2021.cancer-pku.cn/sub-expression.html |
| GEPIA2 | Tang et al. (2019) | http://gepia2.cancer-pku.cn/#index |
| ggplot2 R package | NA | https://ggplot2.tidyverseorg |
| Ggrepel R package | NA | https://cran.r-project.org/web/packages/ggrepel/index.html |
| glmnet R package | Friedman et al. (2010) | https://www.jstatsoft.org/v33/i01/ |
| GSEA-P | Subramanian et al. (2007) | https://www.gsea-msigdb.org/gsea/downloads.jsp |
| Genotype-Tissue Expression (GTEx) | Broad Institute of MIT and Harvard | https://gtexportal.org/home/ |
| ggVennDiagram R package | Gao et al. (2021) | https://cran.r-project.org/web/packages/ggVennDiagram/readme/README.html |
| GOSemSim R package | Yu et al. (2010) | http://bioconductor.org/packages/release/bioc/html/GOSemSim.html |
| ImmuneSigDB | Godec et al. (2016) | http://www.gsea-msigdb.org/gsea/msigdb/genesets.jsp?collection=IMMUNESIGDB |
| maxstat R package | Hothorn et al. (2003)  Lausen et al. (2004) | https://cran.r-project.org/web/packages/maxstat/maxstat.pdf |
| NetworkAnalyst | Xia et al. (2015) | https://www.networkanalyst.ca/ |
| nCounter advanced analysis v2.0.134 | NanoString Technologies | https://www.nanostring.com/products/analysis-solutions/ncounter-advanced-analysis-software/ |
| nSolver v4.0.70 | NanoString Technologies | https://www.nanostring.com/products/analysis-solutions/ncounter-advanced-analysis-software/ |
| PANTHER (v16.0) | Mi et al. (2013)  Thomas et al. (2003) | http://www.pantherdb.org/ |
| pathfindR R package | Ulgen et al. (2019) | https://cran.r-project.org/web/packages/pathfindR/index.html |
| Prism v9.0 | GraphPad | https://www.graphpad.com/scientific-software/prism/ |
| quanTIseq | Finotello et al. (2019) | https://icbi.i-med.ac.at/software/quantiseq/doc/ |
| R v4.0.4 | R Core Team | https://cran.r-project.org/bin/macosx/ |
| SeneQuest | NA | https://senequest.net/ |
| SPSS Statistics v26 | IBM | https://www.ibm.com/uk-en/analytics/spss-statistics-software |
| STRING | Szklarczyk et al. (2019) | http://string-db.org |
| survminer R package | NA | https://cran.r-project.org/web/packages/survminer/survminer.pdf |
| SVA Bioconductor package | Leek et al. (2021) | https://bioconductor.org/packages/sva/ |
| TIDE | Jiang et al. (2018) | http://tide.dfci.harvard.edu/login/ |
| TRRUST | Han et al. (2018) | https://www.grnpedia.org/trrust/ |
| UCell R package | Andreatta et al. (2021) | https://github.com/carmonalab/UCell |
| UCSC Xena | Goldman et al. (2020) | https://xenabrowser.net/datapages/ |

**RESOURCES AVAILABILITY**

**Lead Contacts**

Further information and requests for resources and reagents should be directed to, and will be fulfilled by, the lead contacts, Sergio Rutella and Leo Luznik

**Materials availability**

This study did not generate new unique reagents.

**Data and code availability**

The transcriptomic datasets generated in this study have been deposited on to the GEO repository under accession numbers GSE176100 and GSE178926 and will be publicly available as of the date of publication. The results published here are in part based upon data generated by the TCGA Research Network and by the TARGET initiative, which can be accessed, queried, and visualized through the cBioPortal for Cancer Genomics (<https://www.cbioportal.org/>).

Accessions for gene expression and RNA-sequencing data sets used in this study: newly diagnosed AML GEO: GSE134589 (Vadakekolathu et al., 2020b), newly diagnosed AML TCGA <https://www.cbioportal.org/> (Ley et al., 2013), newly diagnosed AML COG-TARGET <https://www.cbioportal.org/> (Bolouri et al., 2018), newly diagnosed Beat-AML Master Trial <https://www.cbioportal.org/> (Tyner et al., 2018), newly diagnosed AML syn21991338 (Dufva et al., 2020), newly diagnosed AML GEO: GSE76004 (Ng et al., 2016), newly diagnosed AML (German AMLCG 1999 trial) GEO: GSE37642 (Herold et al., 2018; Li et al., 2013), newly diagnosed, chemotherapy-resistant AML GEO: GSE106291 (Herold et al., 2018), untreated cutaneous melanoma TCGA <https://www.cbioportal.org/> (Gao et al., 2013), cutaneous melanoma immunotherapy ENA: PRJEB23709 (Gide et al., 2019), solid tumor immunotherapy GEO: GSE93157 (Prat et al., 2017), tumor microenvironment (TME) classification and functional TME gene signatures <https://science.bostongene.com/tumor-portrait/> (Bagaev et al., 2021).

Codes for reproducibility of data are publicly available or will be available from the corresponding authors upon reasonable request.

**METHOD DETAILS**

**Wet-lab cohorts**

Patient and disease characteristics are detailed in **Table S4**. Primary patient specimens (non-promyelocytic AML) and associated clinical data were obtained *via* informed consent in accordance with the Declaration of Helsinki on research protocols approved by the Ethics Committee of TU Dresden and Studienallianz Leukämie, Germany (EK98032010), and by the Children’s Hospital of Philadelphia (10-007767) and Johns Hopkins University (JHU) Institutional Review Boards. Details on data sources for *in silico* analyses are provided in Supplemental Materials and Methods. The study workflow is illustrated in **Figure 2A**.

**Immunotherapy cohort**

We analyzed NanoString expression profiles of BM samples obtained from 33 elderly patients with chemotherapy-refractory/early relapsed AML on a phase 2 study of AZA+Pembro (clinicaltrials.gov identifier: NCT02845297). Azacitidine was given intravenously at 75 mg/m^2^ daily on days 1 to 7 every 4 weeks, and pembrolizumab was given intravenously at 200 mg on day 8 and every 3 weeks thereafter. Patient and disease characteristics are detailed in **Table S5**.

***In vitro* cytotoxicity assays**

Cytotoxicity of senescent (CD8^+^CD57^+^KLRG1^+^) and non-senescent (CD8^+^CD57^-^KLRG1^-^) T cells against primary AML cells (CD45^low^SSC^int^) was tested *in vitro* using anti–CD33/CD3 and control bi-specific T-cell engager (BiTE) antibody constructs (both provided by Amgen, USA), as previously described (Krupka et al., 2014). Briefly, primary AML samples were sorted into CD8^+^CD57^+^KLRG1^+^ T cells, CD8^+^CD57^-^KLRG1^-^ T cells and AML blasts. T cells were then co-cultured with primary AML blasts (effector/target [E/T] ratio = 1:5) in Iscove’s Modified Dulbecco’s Medium (Life Technologies) supplemented with 15% fetal bovine serum, and 10 ng/ml each of IL-3, SCF, G-CSF, and GM-CSF (all from Life Technologies), for 48 hours. Cells were exposed to either BiTE (10 ng/ml) or cBiTE (10 ng/ml). After 48 hours, T-cell cytotoxicity against CD33^+^CD34^+^ primary AML cells was determined by flow cytometry using the Live/Dead Fixable Yellow Dead Cell Stain Kit (Thermo Fisher Scientific).

**RNA isolation and processing**

RNA was isolated and processed as previously described (Uy et al., 2021; Vadakekolathu et al., 2020b; Wagner et al., 2019). Briefly, 100-150 ng per sample of RNA extracted from BM aspirates were processed on the nCounter FLEX analysis system (NanoString Technologies, Seattle, WA) using the PanCancer Immune Profiling (PCI) panel.

**nCounter data quality control and normalization**

The reporter probe counts, i.e., the number of times the color-coded barcode for that gene is detected, were tabulated in a comma separated value (CSV) format for data analysis with the nSolver software package (version 4.0.62) and nSolver Advanced Analysis module (version 2.0.115; NanoString Technologies, Seattle, WA). The captured transcript counts were normalized to the geometric mean of the housekeeping reference genes included in the assay (n = 40) and the code set’s internal positive controls. Batch effects and other unwanted sources of variation were removed using the Surrogate Variable Analysis (SVA) package in Bioconductor (Leek et al., 2012).

**Validation datasets (AML)**

The Cancer Genome Atlas (TCGA-AML) series consisted of RNA-sequencing data (Illumina HiSeq2000) from 147 adult patients with nonpromyelocytic AML who were enrolled on Cancer and Leukemia Group B treatment protocols 8525, 8923, 9621, 9720, 10201 and 19808. RNA and clinical data were retrieved from cBioPortal for Cancer Genomics (<https://www.cbioportal.org/>) (Ley et al., 2013). Level 3 RSEM-normalized RNASeqV2 data was downloaded and log_2_-transformed prior to analysis. No further pre-processing was applied. For mRNA expression data, cBioPortal for Cancer Genomics computes the relative expression of an individual gene and tumor specimen to the gene’s distribution in all samples that are diploid for the gene in question. The returned value (*z*-score) indicates the number of standard deviations away from the mean of expression in all other tumor samples. To ensure high stringency, a *z*-score threshold of ±2.0 was used in all analyses. Patients had a median age of 60 years, 54% were male, with 12%, 65% and 22% classified as favorable, intermediate, and adverse risk, respectively, based on 2017 European Leukemia Net (ELN) risk stratification by genetics. One hundred thirteen patients (77%) were reported as having received “7+3” cytotoxic induction chemotherapy. The remaining patients were treated with adjunctive therapy in addition to “7+3” or with hypomethylating agents.

The second data series (Beat-AML Master Trial) was retrieved using the VIZOME interface (<http://www.vizome.org/aml/>) and consisted of RNA-sequencing data (Agilent platform) from primary specimens from 281 patients with nonpromyelocytic AML and detailed clinical annotation, including diagnostic information, responses and outcomes, treated on the Beat AML Master Trial (Burd et al., 2020; Tyner et al., 2018).

The third data series, hereafter referred to as the Children’s Oncology Group Therapeutically Applicable Research to Generate Effective Treatments (COG-TARGET) AML series, consisted of RNA-sequencing data (Illumina HiSeq2000) from 145 children, adolescents, and young adults with *de novo* AML enrolled onto biology studies and clinical trials managed through the COG on studies CCG-2961, AAML03P1, or AAML0531 (Bolouri et al., 2018; Farrar et al., 2016).

NanoString immune transcriptomic datasets are available through GEO accession number GSE134589 (n=432 children and adults with newly diagnosed AML) and our previous publication (Vadakekolathu et al., 2020b).

**Validation datasets (melanoma)**

The TCGA Pan-Cancer Atlas series consisted of RNA-sequencing data (Illumina HiSeq2000) from 441 adult patients with untreated primary and/or metastatic melanoma. RNA-sequencing data from 73 patients with melanoma treated with standard-of-care single-agent nivolumab or pembrolizumab (n = 41) or combination anti-PD-1 + anti-CTLA-4 (n = 32) were retrieved through the original publication (Gide et al., 2019) and the Tumor Immune Dysfunction and Exclusion (TIDE) portal (<http://tide.dfci.harvard.edu/login/>) (Jiang et al., 2018). In the original study, responders were defined as individuals with complete response, partial response, or stable disease of greater than 6 months with no progression, and non-responders as progressive disease or stable disease for less than or equal to 6 months before disease progression. NanoString profiling data from 33 patients with advanced solid tumors (n = 9 with melanoma, n = 9 with squamous non-small cell lung carcinoma, n = 12 with non-squamous non-small cell lung carcinoma and n = 3 with head and neck squamous cell carcinoma) treated with anti-PD-1 monotherapy were retrieved through the original publication (GSE9315) (Prat et al., 2017) and the TIDE portal (Jiang et al., 2018).

**Signature calculation**

The relative abundance of immune cell types was computed as previously published (Danaher et al., 2017; Vadakekolathu et al., 2020a). For each sample, immune gene expression scores were calculated as an average (arithmetic mean) of gene expression values for all genes in the signature.

The LSC17 score was computed as the weighted sum of the normalized expression values of the 17 genes included in the signature using the same weights as those provided in the original publication (Ng et al., 2016):

LSC17 score =

(*DNMT3B* × 0.0874) + (*ZBTB46* × −0.0347) + (*NYNRIN* × 0.00865) + (*ARHGAP22* × −0.0138) + (*LAPTM4B* × 0.00582) + (*MMRN1* × 0.0258) + (*DPYSL3* × 0.0284) + (*KIAA0125* × 0.0196) + (*CDK6* × −0.0704) + (*CPXM1* × −0.0258) + (*SOCS2* × 0.0271) + (*SMIM24* × −0.0226) + (*EMP1* × 0.0146) + (*NGFRAP1* × 0.0465) + (*CD34* × 0.0338) + (*AKR1C3* × −0.0402) + (*GPR56* × 0.0501).

**GSEA and leading-edge analysis**

GSEA was performed using the GSEA software v4.1.0 (Broad Institute, Cambridge, MA). A collection of 4,872 gene sets (ImmuneSigDB) derived from 389 published studies of immune cell states and experimental perturbations, both genetic and chemical, was downloaded from <https://www.gsea-msigdb.org/gsea/msigdb/index.jsp> (Godec et al., 2016). Each gene set in the ImmuneSigDB contains either up- or downregulated genes only. The GSEA-p software package was used to extract leading-edge genes that contribute most to the enrichment signal and are shared across the top-ranking ImmuneSigDB gene sets (Subramanian et al., 2005).

**Single-cell RNA sequencing data analysis**

Single-cell RNA sequencing (scRNA) data from eight AML samples were retrieved through the Synapse data repository (https://www.synapse.org/#!Synapse:syn21991338; SynapseID: syn21991338) (Dufva et al., 2020) and analyzed with R v.4.0.2. The R object FIMM_AML_scRNA.Rdata was used without applying any further processing. Plots were generated using Seurat v4.0.1 and custom scripts, which are available from the corresponding authors upon reasonable request.

Single-cell signature scores were estimated with the function AddModuleScore_UCell (UCell v1.0.0 package, available on GitHub at https://github.com/carmonalab/UCell), using default settings. This function is based on the Mann-Whitney *U* statistic and calculates scores based on the relative ranking of genes for individual cells. Gene lists generated in this study are provided in **Table S2**.

**Statistical testing**

Descriptive statistics included calculation of median, inter-quartile ranges and proportions to summarize study outcomes. Comparisons were performed with the Mann-Whitney *U* test for paired or unpaired data (two-sided), as appropriate, or with the ANOVA with correction for multiple hypothesis testing. Given the potentially large number of parameters with high correlation and in order to prevent overfitting, we used the Least Absolute Shrinkage and Selection Operator (LASSO) regularization technique for variable reduction (*glmnet* package in R) (Tibshirani, 1997). Ten-fold cross-validation was used to select the optimal regularization parameter. Genes with nonzero coefficients were selected as predictive of the outcome variable (patient survival).

Overall survival was computed from the date of diagnosis to the date of death. Relapse-free survival was measured from the date of first complete remission to the date of relapse or death. Subjects lost to follow-up were censored at their date of last known contact. Kaplan-Meier survival plots were generated using the *survminer* package in R and the log-rank (Mantel-Cox) test was used to compare survival distributions. The *P* values were adjusted for multiple hypothesis testing using the Benjamini-Hochberg procedure. The alpha value for all comparisons was 0.05. IBM SPSS Statistics (version 27), R (version 4.1.1) and GraphPad Prism (version 9.3.1) were used for statistical analyses.
